# Running therapy improves clinical symptoms and reorganizes dynamic brain networks in affective disorders

**DOI:** 10.64898/2026.01.30.26345203

**Authors:** Julian Gaviria Lopez, Guido Van Wingen, Chris Vriend, Laura K.M. Han, Jennifer Labus, Gitte M. Knudsen, Brenda W.J.H. Penninx

**Author notes:** Correspondence Julian Gaviria Lopez.

## Abstract

**Background:** Exercise therapy reduces depressive and anxiety symptoms, but its neural mechanisms are not fully understood. We examined whether and how running therapy reorganizes dynamic brain functional connectivity in affective disorders.

**Methods:** At baseline, resting-state fMRI was collected from 66 healthy controls and 50 individuals with affective disorders. Co-activation patterns analyses (CAPs) identified recurring whole-brain network states characterized by spatial patterns of regional co-activation/codeactivation patterns and their temporal occurrence rates. We compared CAPs between groups at baseline. Participants with affective disorders then received 16 weeks of running therapy or antidepressant treatment. We examined: (1) treatment-induced changes in brain CAPs and clinical symptoms, (2) brain-symptom associations at baseline versus post-treatment, and (3) associations between network reorganization and symptom improvement.

**Results:** At baseline, individuals with affective disorders showed fewer occurrences of the visual-somatomotor-subcortical network state (VS-SCCAP) than controls (F=5.4, P=0.02, η²=0.04). Running therapy significantly altered the temporal dynamics of two brain systems: the default mode (DM_CAP_: β = -0.88, P = 0.006, d =- 0.88) and VS-SC_CAP_ (β = 0.87, P = 0.006, d = 0.85). These reorganizations were accompanied by significant improvements in depressive and anxiety symptoms (IDS: β = -1.23, P < 0.001, d = -1.15; BAI: β = - 0.98, P = 0.008, d = -0.93). DM_CAP_-symptom coupling changed significantly from baseline to post-treatment (ΔRHO=-0.48, Z≈-2.0, P<0.05).

**Conclusions:** Running therapy altered dynamic brain networks in association with clinical symptom improvement. These findings provide neurobiological evidence for exercise-induced therapeutic effects through transient brain-state reorganization, demonstrating the utility of dynamic connectivity approaches for characterizing neural mechanisms in affective disorders.

## Introduction

Affective disorders involve dysfunction across multiple neural systems (Craske et al., 2017; Otte et al., 2016). However, the brain mechanisms underlying therapeutic interventions remain unclear. Running therapy has emerged as an effective alternative, demonstrating clinical improvements comparable to selective serotonin reuptake inhibitors (SSRIs) for mild-moderate depression (Blumenthal et al., 2007; Knapen et al., 2015; Ravindran et al., 2016; Pearce et al., 2022), with benefits for metabolic and inflammatory outcomes in patients with immunometabolic depression features (Vreijling et al., 2025). The MOTAR study (Lever-van Milligen et al., 2019) directly compared 16 weeks of running therapy versus antidepressant treatment (escitalopram or sertraline) in 141 individuals with depression and/or anxiety disorders, revealing comparable mental health remission rates (antidepressants: 44.8%; running: 43.3%) but significantly superior physical health outcomes in the running group, including improvements in weight, waist circumference, blood pressure, heart rate, and heart rate variability (Verhoeven et al., 2023). However, the neural mechanisms through which exercise ameliorates affective symptoms remain poorly characterized (Firth et al., 2018). Previous neuroimaging research in affective disorders has revealed alterations within and between the brain’s default mode network (DMN), salience network (SN), and frontoparietal network (FPN) (Kaiser et al., 2015; Mulders et al., 2015; Northoff, 2016). Specifically, regarding the neural substrates underlying the effects of running therapy on affective disorders, Vriend et al. (2025), using the MOTAR MRI subsample (Figure 1), found no differences in brain network connectivity between controls and individuals with affective disorders at baseline, nor pre-post treatment changes following running therapy, despite significant clinical improvement in the MRI subsample and full MOTAR cohort (Verhoeven et al., 2023).

**Figure 1.**
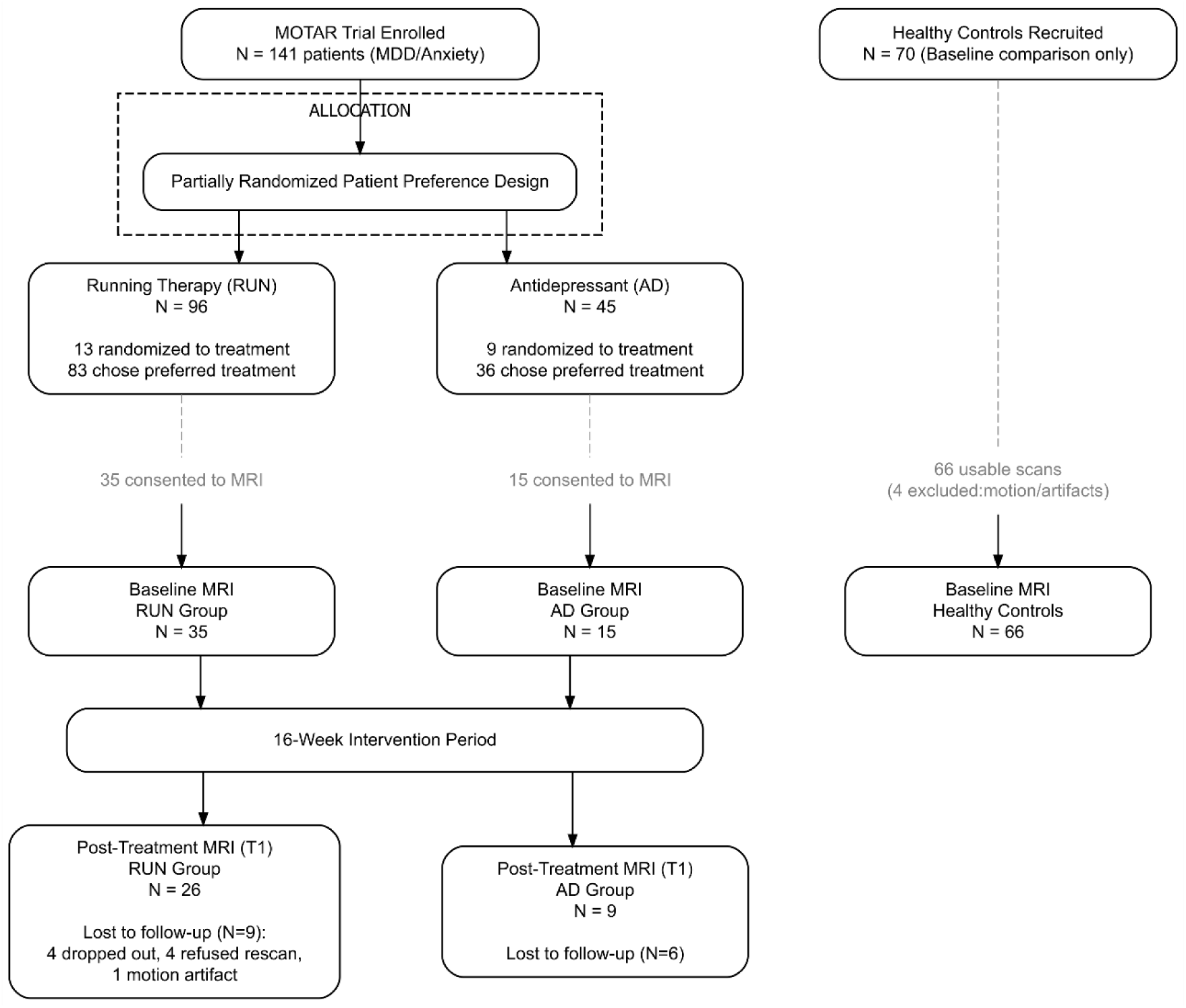
CONSORT flow diagram for the MOTAR FMRI substudy. Individuals suffering from affective disorders semi-randomized to running (RUN, n=96) or antidepressants (AD, n=45), plus healthy controls (CTR, n=70). Baseline/follow-up fMRI completed by: RUN 35/26, AD 15/9, CTR 66/—.

These previous neuroimaging studies have employed methods that assume constancy and homogeneity in the functional brain (Haak et al., 2018). Specifically, their functional connectivity (FC) metrics average the brain’s BOLD (blood-oxygenated) response across the entire scanning session and rely on functional atlases that compartmentalize the brain into patches of assumed piecewise homogeneous connectivity. These assumptions may obscure transient network reconfigurations that could be critical to understanding the effects of treatment on affective disorders. Notably, the null findings reported by Vriend et al. (2025) on static FC underscore the need for dynamic functional connectivity (dFC) approaches that capture transient brain network engagement in response to aerobic exercise. One method to characterize dFC is co-activation pattern (CAPs) analysis, a data-driven approach that identifies recurring whole-brain states and their temporal occurrence rates (Liu and Duyn, 2013; Bolton et al., 2020). This approach enabled us to define dFC configurations between regions (i.e., CAPs) at voxel-wise spatial resolution and fMRI-frame temporal resolution, which traditional methods based on a priori compartmentalization of brain areas (i.e., functional atlas) do not capture. The CAPs approach has proven beneficial for studying brain correlates of affective processing (Gaviria et al., 2021; Morgenroth et al., 2023), the brain substrates underlying the interplay between emotions and cognition (Gaviria et al., 2020), and the dynamics of amygdala connectivity in bipolar disorders across mood states (Rey et al., 2021; Saccaro et al., 2023).

The present study applies dFC to resting-state fMRI data from the MOTAR MRI substudy (Figure 1) to test three complementary hypotheses: I) Individuals with affective disorders would exhibit altered coactivation patterns (i.e., CAPs) compared with healthy controls at baseline, reflecting disrupted temporal dynamics of large-scale network engagement. II) Running therapy would induce treatment-specific brain network reorganization distinct from SSRI treatment, as evidenced by differential longitudinal trajectories. III) Brain CAPs that respond to treatment would show altered brain-symptom coupling from baseline to post-treatment. The magnitude of brain network reorganization would predict individual symptom improvement. By characterizing transient brain-wide network states modulated by treatment and defining their relationships with clinical outcomes over time, we aimed to elucidate the neural mechanisms through which running therapy ameliorates affective disorders.

## 2. METHODS

### 2.1. Participants

Eligibility criteria for MOTAR are described in detail in the protocol paper (Lever-van Milligen et al., 2019). Briefly, individuals between 18 and 70 years diagnosed with a current major depressive disorder (MDD), social phobia, generalized anxiety disorder, panic disorder, or agoraphobia according to DSM-IV criteria were recruited between July 2012 and July 2019. Diagnostic criteria were verified by a trained researcher using the Composite International Diagnostic Interview (CIDI). Exclusion criteria included antidepressant use within the past two weeks, current use of psychotropic medication other than stable benzodiazepine use, exercising more than once a week, having a psychiatric diagnosis other than depression or anxiety disorder, acute suicidal risk, somatic contraindications to running therapy or treatment with antidepressants, or pregnancy. Severe somatic contraindications that might interfere with safe participation in the running sessions were discussed with participants’ physicians before enrollment. Additionally, healthy participants without any history of psychiatric disorders were recruited as a comparison group for baseline case-control analyses. For the MRI sub-study, participants needed to be free from metallic implants and claustrophobia. The MOTAR trial enrolled 141 participants with affective disorders (96 on running therapy, 45 on SSRI antidepressants); 50 of them met criteria and agreed to participate in the MRI substudy (35 on running therapy, 15 on SSRI). Combined with 70 healthy controls (4 were discarded due to MRI issues), the baseline (T0) sample included 116 participants: 66 controls (36 male, 30 female) and 50 with affective disorders (28 male, 22 female). Post-intervention (T1, Week 16), MRI data were collected from 39 participants (26 on running therapy, 9 on SSRI). A detailed CONSORT flow diagram for participant enrollment and retention in the MRI substudy is provided in Figure 1. All participants gave informed consent. The study was approved by the medical ethics committee of VU University Medical Center and conducted in accordance with the Declaration of Helsinki. The trial was registered prospectively in the Netherlands Trial Registry (NTR3460).

### 2.2. Study Design

The MOTAR study employed a partially randomized patient preference design in which 141 individuals with major depressive disorder and/or anxiety disorder received 16 weeks of treatment. Participants were either randomized or allowed to choose between two interventions: (1) antidepressant medication (hereafter “AD” group) or (2) outdoor running therapy (hereafter “RUN” group). Antidepressant (AD) group: participants received escitalopram (10–20 mg/day) or sertraline (50–200 mg/day) if escitalopram was poorly tolerated or ineffective. All participants were medication-free for at least 2 weeks before enrollment, though stable benzodiazepine use was permitted. Running (RUN) group: Participants completed at least two supervised outdoor running sessions per week (45 minutes each). Each session consisted of a 10-minute warm-up, a 30-minute progressive-intensity jog (50–70% heart rate reserve in weeks 1–4; 70–85% in weeks 5–16), and a 5-minute cool-down. Three group sessions were offered weekly, plus one optional individual session. Adherence was monitored via attendance records and continuous heart rate monitoring. The AD group served as an active treatment comparison to contextualize the specificity of running-induced brain network trajectories (Figure 5B), but it was not included in the primary within-group longitudinal analyses (Figure 5A, C, D) due to a smaller sample size (N=15 at T0; N=9 at T1) and our specific focus on mechanisms underlying running therapy.

**Figure 2.**
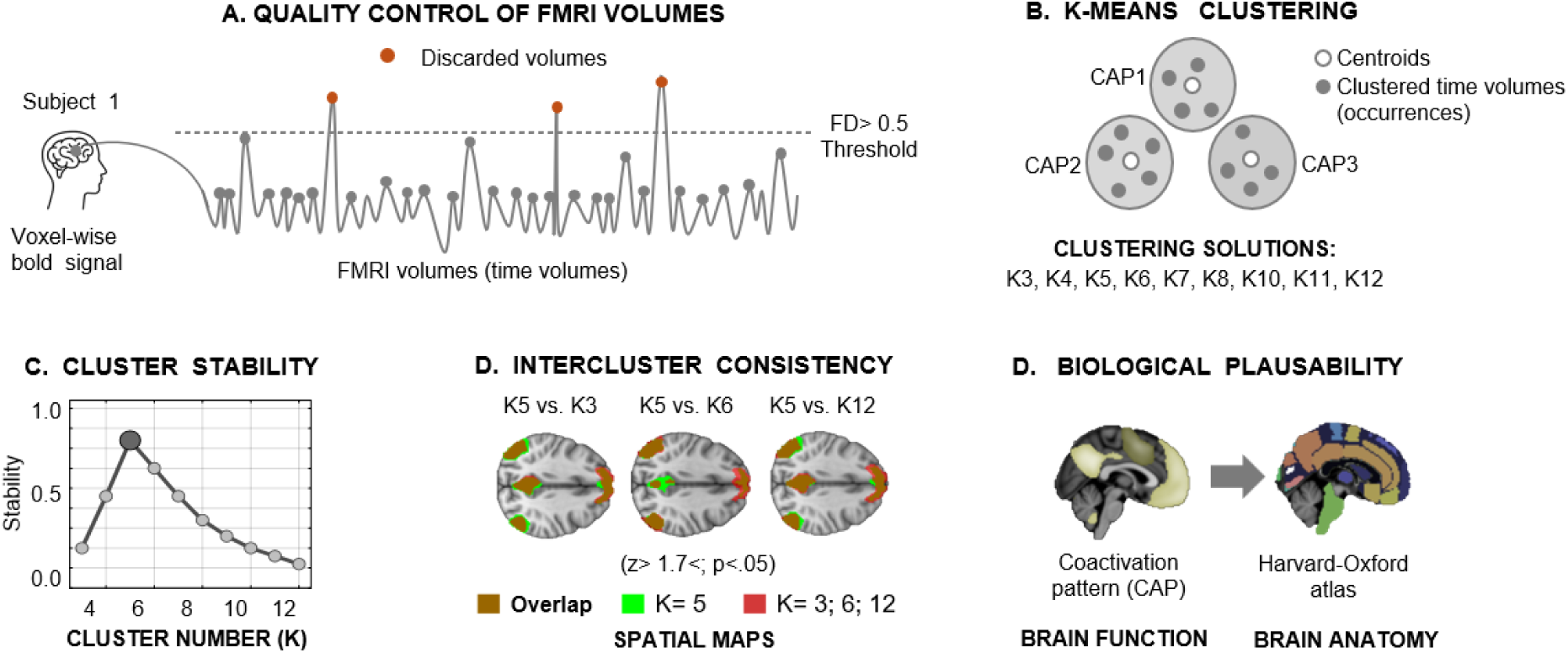
CAP Analysis Pipeline for fMRI Data in the MOTAR Study. **A.** Quality control of FMRI volumes for CAP analysis. Voxel-wise BOLD signal fluctuations are computed across all fMRI volumes. Volumes contaminated by head motion (framewise displacement > 0.5 mm, orange dots) are excluded. Retained volumes (gray dots) are submitted to k-means clustering based on their spatial similarity (Liu et al., 2013). (Panel B). Peaks in the time series indicate moments when specific CAPs are active. For example, if CAP1 appears in 600 of 2,000 volumes, its occurrence rate is 30%. Higher occurrence rates indicate more frequent network engagement. **B.** K-means clustering identified three distinct brain states representing recurrent whole-brain co-activation patterns. Three complementary metrics identified the optimal number of brain clusters (i.e., CAPs): **C.** Cluster stability assessment, **D.** Intercluster consistency, **E.** Biological plausibility.

**Figure 4.**
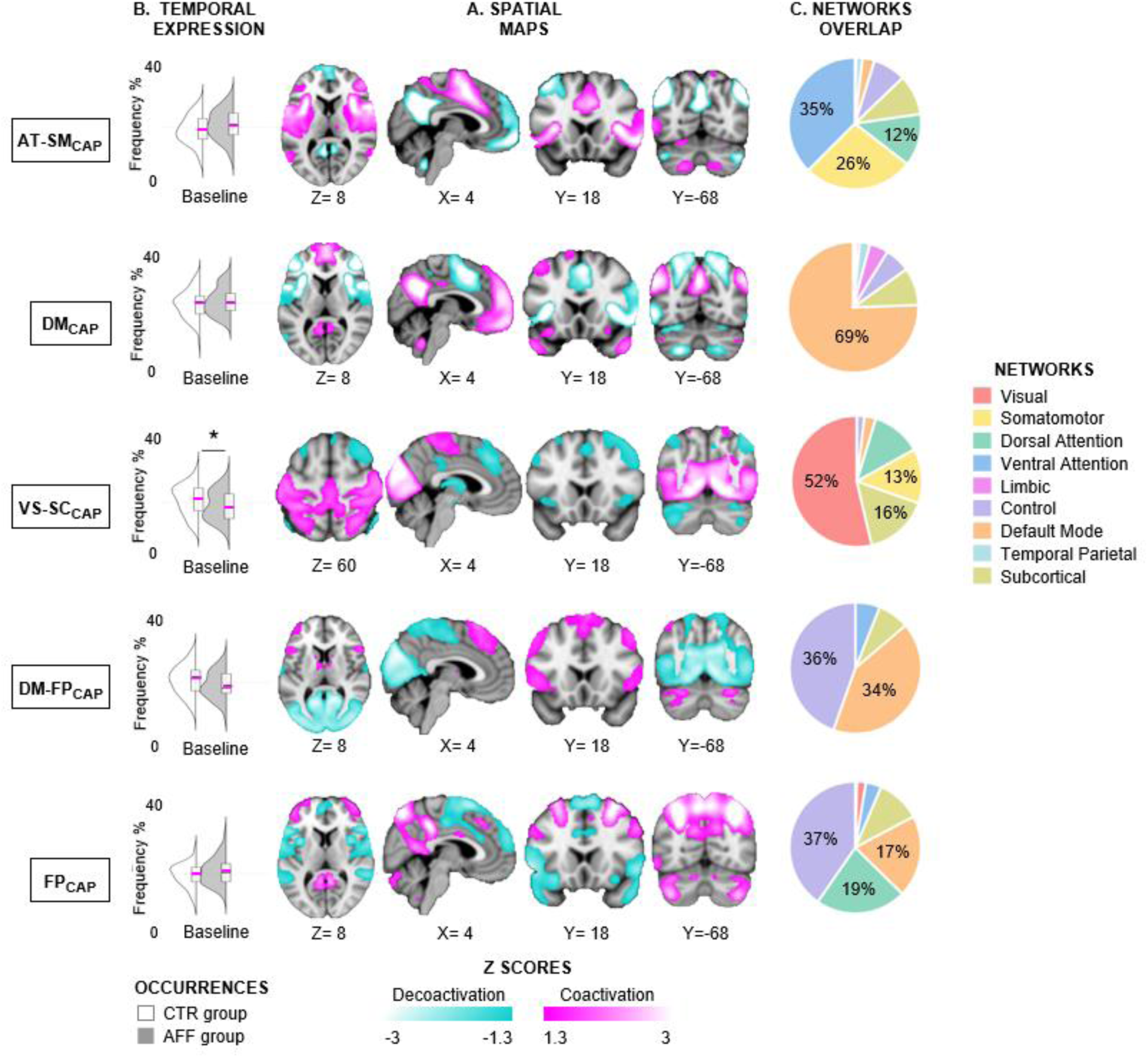
Spatial, temporal, and network characteristics of CAPs. **A.** Spatial maps (magenta = coactivation; cyan = codeactivation) for five CAPs. **B.** CAP occurrence frequency at baseline (controls vs. patients). Violin plots show distribution; asterisks indicate significant group differences (*p < 0.05) after adjusting for age and sex using ANCOVA. Full results are described in Table 2. **C.** Network overlap with canonical brain networks (Schaefer-400 atlas). Five CAPs identified: AT-SMCAP, DMCAP, VS-SCCAP, DM-FPCAP, and FPCAP. Network colors are consistent across all panels.

**Figure 5.**
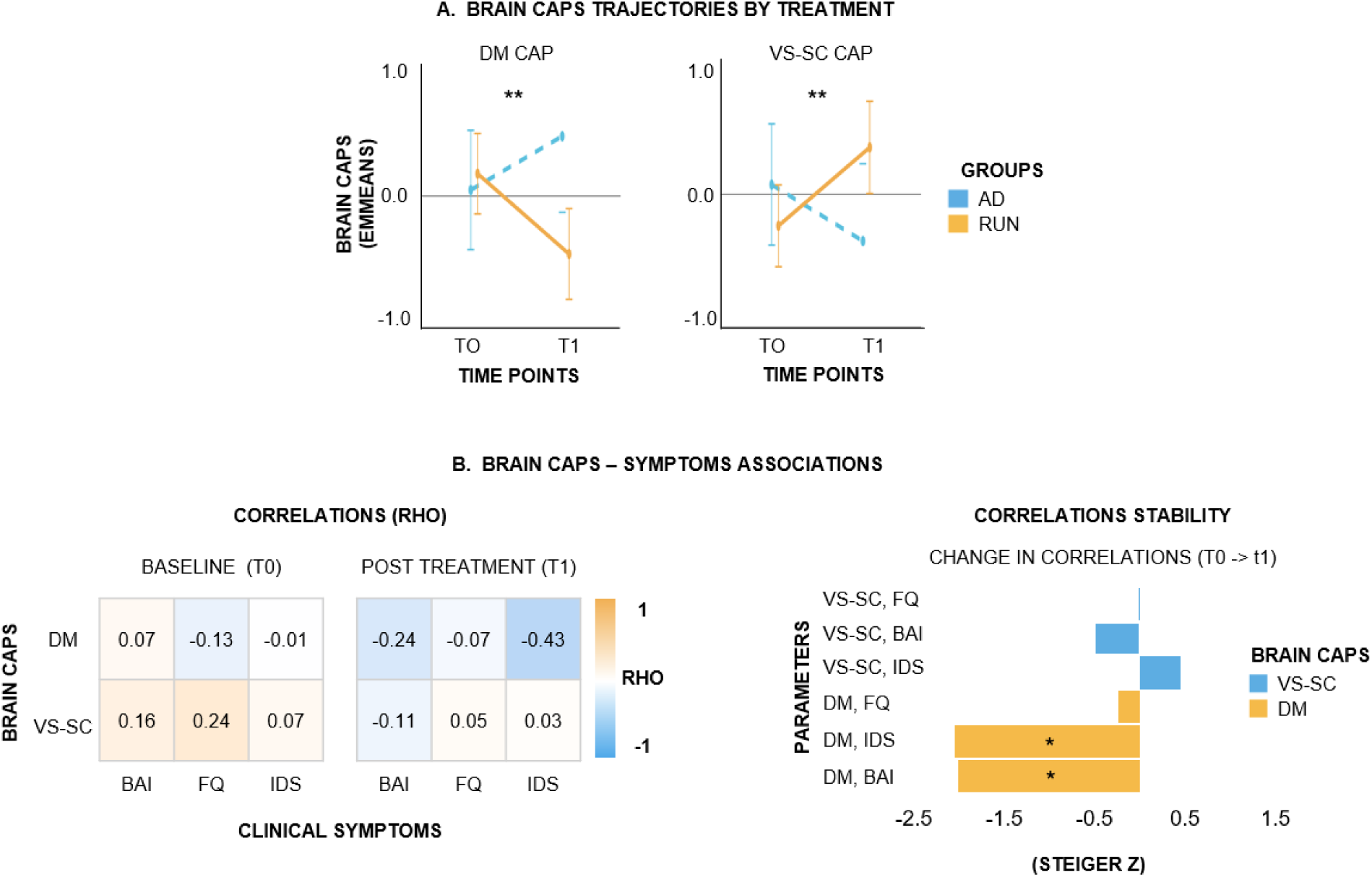
Running therapy reorganizes brain networks and reconfigures brain-symptom coupling. **A.** VISIT × TREATMENT interactions revealed treatment-specific CAP trajectories (η²p = 0.13, p = 0.01 for both CAPs): running therapy (RUN) produced large within-group changes (DMCAP: d = −0.88; VS-SCCAP: d = 0.85, both p < 0.001), while antidepressants (AD) showed negligible changes (|d| ≤ 0.61, p ≥ 0.18; Table S17). Y-axis shows estimated marginal means (emmeans) of z-scored CAP occurrence rates, representing the predicted frequency of each brain state. **B.** CAP-symptom correlations (Spearman’s ρ) were predominantly non-significant at both baseline (T0) and post-treatment (T1). Correlation stability analysis (Steiger’s Z-test, permutation-validated) revealed that DMCAP-symptom relationships changed significantly between timepoints (IDS: Z = −2.00, p = 0.05; BAI: Z = −2.05, p = 0.04), shifting from weak positive/null at baseline to moderate negative post-treatment. In contrast, all other correlations remained stable (|Z| < 0.50, p > 0.60), indicating that network reorganization occurred with selective reconfiguration of brain-symptoms coupling. Tables S15-S17. FQ: Fear Questionnaire; IDS: Inventory of Depressive Symptomatology; BAI: Beck Anxiety Inventory.

### 2.3. Clinical measurements

At the day of MRI scanning, we administered the Inventory of Depressive Symptomatology (IDS; 30 items) (Rush et al., 1996), the Beck Anxiety Inventory (BAI; 21 items) (Beck et al., 1988), and Fear Questionnaire (Marks and Mathews, 1979), to measure the severity of depressive, anxiety and phobia symptoms, respectively.

### 2.4 MRI Acquisition

MRI scans were acquired at the Spinoza Centre for Neuroimaging (Amsterdam, the Netherlands) using a Philips 3T Achieva MR scanner (Philips Healthcare, Best, the Netherlands) with a 32-channel head coil. Structural images were acquired using 3D T1-weighted Turbo Field Echo (TR = 8.1 ms, TE = 3.7 ms, flip angle = 8°, 1 mm³ isotropic voxels). Resting-state fMRI data (8 minutes, eyes closed) were acquired using T2*-weighted echo-planar imaging (TR = 2300 ms, TE = 28 ms, flip angle = 76°, 3.0 × 3.0 mm² in-plane resolution, 37 sequentially ascending axial slices with 3 mm thickness and 0.3 mm gap, 210 volumes). Identical protocols were used at all timepoints. See Supplementary Methods for additional details on acquisition and preprocessing.

### 2.5. Dynamic functional connectivity analysis

#### 2.5.1. Co-activation patterns (CAPs)

To capture time-varying brain dynamics, we calculated co-activation patterns, a data-driven, seed-free method for identifying recurring whole-brain states during resting-state fMRI (Bolton et al., 2020; Liu and Duyn, 2013). Unlike static measures that average connectivity across the entire scan, CAP analysis describes transient configurations of coordinated activation and deactivation across distributed brain regions, offering insights into the temporal dynamics of large-scale network engagement. The **CAP maps** (i.e., the spatial metric) show which brain regions co-activate (or codeactivate), while temporal metrics measure when and how often these patterns occur. We calculated **CAP occurrences**—the percentage of fMRI time frames during which each brain state was present. For instance, if CAPx appeared in 60 of 200 total frames, its occurrence would be 30%. Higher occurrence rates suggest a network state appears more frequently during the fMRI scan; lower rates indicate less frequent engagement (see the illustration of this metric in Figure 2A). This metric of temporal variability was used to study both baseline group differences and treatment-related changes in the dynamic expression of brain CAPs. A detailed description of the CAPs generation and validation is provided in the Supplementary methods.

### 2.6 Statistical Analysis

#### 2.6.1. Demographic Covariate Assessment

To assess the impact of demographics on CAP expression, we examined associations between age, sex, and CAP temporal occurrence (percentage of fMRI time points assigned to each CAP) in two samples: (1) the overall baseline sample (N=116: 66 controls), and (2) stratified by diagnostic group. Associations with age were quantified using Pearson correlations (*r* with 95% confidence intervals). Sex differences were evaluated using Mann-Whitney U tests (non-parametric) with rank-biserial correlation (r_rb_) effect sizes.

#### 2.6.2. Baseline comparisons between controls and individuals with affective disorders

To compare the temporal occurrence rates of CAPs between controls and individuals with affective disorders at baseline (T0), we employed independent samples t-tests and reported Cohen’s d effect sizes. To verify robustness and address potential demographic confounding, we re-analyzed differences using Analysis of Covariance (ANCOVA), controlling for age and sex, despite minimal demographic main effects (Supplementary Figure S2; all |r|<0.20, P>0.15). ANCOVA models used Type II sums of squares, with partial eta-squared (η²p) quantifying effect sizes.

#### 2.6.3. Treatment Effects on Brain CAPs and Clinical Symptoms

##### 2.6.3.1. Changes in brain CAPs induced by treatment

Linear mixed models (LMMs) compared running therapy (RUN group: N=35→26) and SSRI treatment (AD group: N=15→9) using the formula: Outcome (CAPx) ∼ VISIT × TREATMENT + sex + age + (1|SUBJECT). The VISIT × TREATMENT interaction tested whether T0-to-T1 changes differ between groups. Models used REML estimation with Kenward-Roger degrees of freedom. Given the smaller AD group size (N=9 at T1), we implemented: (1) parametric bootstrap validation (N=5,000 iterations) to generate 95% CIs for interaction effects; (2) post-hoc power analysis to determine minimum detectable effect sizes (MDE) for 80% power; and (3) standardized effect size reporting (Cohen’s d, partial η²) to emphasize magnitude over significance. Effects were normalized using z-scores. Moreover, the FDR correction procedure was applied to account for multiple comparisons across CAPs. Model diagnostics were used to evaluate model convergence, residual distributions, and the adequacy of the random-effects structure for this dataset. This analytic approach allowed us to: (1) examine within-group changes over time for each treatment separately, (2) compare between-group trajectories—i.e., whether the magnitude and direction of T0-to-T1 changes differed between running therapy (RUN) and SSRI antidepressant (AD) groups—and (3) identify treatment-specific brain network reorganization patterns. Notably, assessing treatment effects through VISIT × TREATMENT interaction analyses was necessary because comparing separate within-group analyses (e.g., testing whether each group changed significantly from baseline) does not constitute a valid test of differential effects (Nieuwenhuis et al., 2011). Moreover, the interaction term isolates treatment effects from temporal confounds (e.g., test-retest effects) that could otherwise confound single-group pre-post comparisons.

#### 2.6.4. Brain-Symptom Associations

To examine whether treatment altered brain-symptom coupling, we analyzed Spearman’s rank correlations between CAP occurrence rates and clinical symptom scores at baseline (T0) and post-treatment (T1). Analyses were restricted to CAPs demonstrating significant VISIT × TREATMENT interactions in Stage 1 LMM analyses, justified by: (1) limited sample size necessitating targeted hypothesis testing, and (2) the biological premise that unchanged variables cannot mediate treatment effects. **Temporal correlation stability testing**. To test whether correlation strengths changed between T0 and T1, we applied Steiger’s Z-test (Steiger, 1980), which accounts for autocorrelation in dependent correlation comparisons. Robustness was confirmed via non-parametric permutation testing (5,000 iterations), in which timepoint labels were randomly shuffled to generate null distributions of correlation differences. **Change-change associations**. We also examined whether the magnitude of brain network reorganization was associated with symptom improvement by computing change scores (Δ = T1 − T0) for treatment-responsive CAPs and clinical measures, then correlating ΔCAP with ΔSymptom values using Spearman’s rank correlation. The antidepressant (AD) group was excluded from brain-symptom correlation analyses due to our primary interest in the brain systems underlying the effects of physical exercise on affective disorders, and the insufficient AD-paired data (N=9).

##### Statistical Analyses Summary

First, baseline analyses quantified differences in CAP occurrence rates between controls and individuals with affective disorders. Second, treatment effects were evaluated via linear mixed models comparing running therapy and SSRI treatment, with the VISIT × TREATMENT interaction testing trajectory differences between groups. Third, brain-symptom associations were examined using non-parametric correlations restricted to treatment-responsive CAPs. Post-hoc tests assessed temporal stability (i.e., whether correlation strengths differed between T0 and T1) using Steiger’s Z-test and permutation testing. Change-change associations (corr(ΔCAP, ΔSymptom)) assessed whether brain reorganization magnitude predicted symptom improvement. Multiple robustness measures were implemented across all analytical steps to ensure the reliability of parameter estimates (including demographic covariate control, bootstrap validation, post-hoc power analysis, permutation testing, standardized effect sizes, FDR correction, and model diagnostics.

## 3. RESULTS

### 3.1. Demographic and clinical characteristics

In line with Vriend et al., 50 participants with a depressive or anxiety disorder and 66 healthy controls were included in our analyses at baseline (Figure 1). Of the 50 individuals with an affective disorder, 20 % had major depressive disorder, 28% had an anxiety disorder (social phobia, panic disorder, agoraphobia, or generalized anxiety disorder), and 52 % had comorbid depression and anxiety disorder (Table 1). As expected, individuals with an affective disorder scored significantly higher on the depression (IDS), anxiety (BAI), and fear (FQ) questionnaires compared with healthy controls (all P < 0.001). Groups were well matched on age and sex, but not on education (U = 1172.5, P < 0.01). The results are presented in Table 1.

**Table 1.** Demographic and clinical characteristics. Data are presented as mean (SD) unless otherwise indicated. Sample characteristics and intervention details have been previously described (Vriend et al., 2025; Verhoeven et al., 2023). Case-control statistics compare healthy controls (n=66) versus individuals with affective disorders (N=50) using Mann-Whitney U tests (continuous) and chi-square tests (categorical). The running therapy subsample (N=26) represents completers analyzed in the present neuroimaging study. Treatment assignment: 81% preferred, 19% randomized. *Multiple diagnoses possible (comorbidity). Compliance: Percentage (%) of patients that followed the entire running therapy according to protocol.. IDS, Inventory of Depressive Symptomatology; BAI, Beck Anxiety Inventory; FEAR, Fear Questionnaire; SSRI, selective serotonin reuptake inhibitor.

| Variable | Healthy controls<br>(n=66) | Individuals with<br>affective disorder<br>(N=50) | Case-control<br>Statistics | Running therapy<br>sample (N=26) |
| --- | --- | --- | --- | --- |
| Sex (N (%)) |  |  |  |  |
| Male | 36 (55%) | 22 (46%) | $\chi^2_{(1)} = 0.88, P = 0.35$ | 14 (54%) |
| Female | 30 (45%) | 28 (54%) |  | 12 (46%) |
| Age (years) | 39.5 (14.0) | 35.5 (11.5) | U = 1877, P = 0.21 | 38.4 (12.6) |
| [range] |  |  |  | [22 – 66] |
| Education (years) | 13.3 (3.3) | 11.7 (3.1) | U = 1172.5P < 0.01 | 12.1 (2.8) |
| IDS | 0.5 (0.7) | 16.2 (13.9) | U = 18 P < 0.001 | 37.0 (10.5) |
| BAI | 1.5 (2.0) | 22.1 (12.2) | U = 24 P < 0.001 | 21.3 (10.6) |
| FEAR | 8.8 (9.7) | 40.3 (26.8) | U = 364 P < 0.001 | 35 (19.4) |
| Diagnosis* |  |  |  |  |
| Major depressive disorder | – | 36 (72%) | – | 16 (64%) |
| Social Phobia | – | 21 (42%) | – | 8 (32%) |
| Panic disorder | – | 21 (42%) | – | 7 (28%) |
| Agoraphobia | – | 13 (26%) | – | 5 (20%) |
| Generalized Anxiety Disorder | – | 15 (30%) | – | 9 (36%) |
| Treatment (running therapy / SSRI) | – | 35 / 15 | – | 25 |
| Preferred or randomized treatment (%) | – | – | – | 81% / 19% |
| Compliance | – | – | – | 57.1% |

### 3.2. Co-activation patterns (CAPs) identification and validation

In our dFC analysis, co-activation patterns were computed using k-means clustering on concatenated resting-state fMRI frames from baseline and post-treatment timepoints (T0, N= 116; T1, N= 35; Figure 2A-B). This data-driven approach captured dynamic functional coordination patterns that fluctuate over time, circumventing limitations of static connectivity methods and a priori anatomical parcellations. A systematic evaluation of clustering solutions for k=2 to k=17, using three complementary criteria—clustering stability (consensus index across 100 initializations), inter-cluster consistency (normalized mutual information), and biological plausibility (spatial overlap with Harvard-Oxford atlases)—identified k=5 as the optimal solution. The k=5 solution showed peak stability (consensus >0.22), strong correspondence between coarser and finer solutions (NMI >0.7), and superior neuroanatomical alignment (median Dice ∼0.80). A graphical summary of the results is provided in Figure S1A-D. Cross-validation across three independent clustering scenarios (all participants, CTR group only, AFF group only) confirmed robustness, with high spatial similarity (Dice >94%, Jaccard >88%) and temporal similarity (Pearson r >0.9, concordance >0.7), establishing that the five identified CAPs represent generalizable brain states independent of sample composition (see Supplementary Figure S2 A-C) for details in the validation procedure).

### 3.3. The spatial representation of brain CAPs

The first brain CAP exhibited coordinated engagement of sensorimotor networks (supplementary motor area, supramarginal gyrus, rolandic operculum, pre-/postcentral gyri), salience/ventral attention networks (bilateral insula, mid-cingulate), temporoparietal regions (superior temporal gyrus, Heschl’s gyrus), and frontoparietal control regions (inferior/middle frontal gyrus, inferior parietal lobule). We defined CAP1 as Attention-Sensorimotor CAP (i.e., **AT-SM_CAP_**) because areas of the sensorimotor networks and salience/attention networks together comprise 73% of co-activation (Figure 3C, Table S1). Concomitant suppression (i.e., codeactivation) predominantly encompasses default mode regions (precuneus, angular gyrus, medial prefrontal cortex, posterior cingulate), frontoparietal control regions (superior frontal gyrus, middle frontal gyrus), visual cortex, and the cerebellum. See full description in Table S2.

Brain CAP2 primarily shows activation of default mode subsystems, including the anterior default mode (medial prefrontal cortex, orbitofrontal cortex, rectus gyrus), the posterior default mode (precuneus, angular gyrus, posterior cingulate), and the lateral temporal default mode regions (middle temporal gyrus). Additional co-activation involves control, limbic, and visual areas (specifically, the cuneus and middle occipital gyrus). Because of the high overlap (69%) with brain regions generally linked to the default mode network, we labelled CAP2 as **DM_CAP_** The reciprocal codeactivation profile included frontoparietal control areas (inferior parietal lobule, dorsolateral prefrontal cortex, inferior frontal gyrus), sensorimotor networks (supramarginal gyrus, supplementary motor area, precentral and postcentral gyri, rolandic operculum), salience/ventral attention networks (bilateral insula, mid-cingulate), and dorsal attention networks (superior parietal lobule). See full description in Figure 3, Tables S3 and S4.

CAP3 included co-activation of the cortical central visual and peripheral visual areas with regions associated with sensorimotor networks, specifically the precentral and postcentral gyri, and the paracentral lobule. Additionally, this coactivation pattern included the dorsal attention system, particularly within the superior parietal lobule, as well as subcortical areas including the middle and inferior temporal gyri and the precuneus. This configuration, incorporating visual, sensorimotor, and subcortical networks, was designated as **VS-SC_CAP_**. Refer to Figure 3 and Table S5 for further illustration. Synchronized codeactive systems identified in VS-SC_CAP_ comprised brain regions pertinent to frontoparietal control networks, such as the inferior parietal lobule, middle frontal gyrus, and inferior frontal opercularis, as well as components of the default mode network, including the angular gyrus, medial prefrontal cortex, and inferior parietal lobule. Furthermore, the thalamic nuclei involved the ventral lateral, mediodorsal, ventral anterior, anterior ventral, lateral posterior, intralaminar, pulvinar, and medial dorsal lateral structures. See full description in Tables S5 and S6.

CAP4 was designated as **DM-FP_CAP_** due to its co-activation within the default mode and frontoparietal control networks, accounting for 70% overlap with aprioristic canonical parcellations (Schaefer et al., 2018). Specifically, in the medial and dorsolateral prefrontal cortex, as well as the angular and inferior parietal regions. This co-activation occurred concurrently with extensive co-deactivation observed in visual areas (∼70%; including the occipital cortex, calcarine, and cuneus) and sensorimotor regions (∼28%; encompassing the central gyri and supplementary motor area) (see Figure 3 and Supplementary Tables S7–S8).

CAP5 showed distributed activation across frontoparietal regions (dorsolateral prefrontal cortex, inferior parietal lobule, inferior frontal gyrus), dorsal attention areas (superior parietal lobule, intraparietal sulcus, angular gyrus), and default mode network components (precuneus, posterior cingulate, lateral temporal cortex). Here, they are referred to as the frontoparietal **FP_CAP_** (Figure 4). . Concomitant suppression (i.e., codeactivation) encompassed the medial prefrontal cortex, lateral temporal cortex, temporal pole, angular, precuneus, inferior temporal, sensorimotor regions (supplementary motor area, supramarginal gyrus, rolandic operculum, pre-/postcentral, paracentral), and regions linked to the limbic network (orbitofrontal cortex, lateral orbitofrontal posterior, bilateral insula). See full description in Tables S9 and S10.

**Table 2.** Group differences in dynamic brain network expression. Co-activation patterns (CAP) temporal occurrence at baseline (T0) compared to individuals with affective disorders (MDD, N=50) and healthy controls (CTR, n=66). Left columns: Cohen’s d effect sizes with 95% confidence intervals. Right columns: ANCOVA results controlling for age and sex (Effect size: partial η²). Positive d indicates greater occurrence in MDD. Visual-Subcortical (VS-SC) and Default Mode (DM) CAPs exhibited higher expression in MDD after covariate adjustment. FP, Frontoparietal; AT-SM, Attention-Sensorimotor.

| T-TEST |  |  |  |  | ANCOVA |  |  |  |
| --- | --- | --- | --- | --- | --- | --- | --- | --- |
| CAP | P | d | CI_lower | CI_upper | F | P | $\eta^2$ | $\beta$ |
| FP | 0.07 | -0.34 | -0.71 | 0.03 | 3.9 | 0.05 | 0.03 | 0.37 |
| DM | 0.28 | -0.20 | -0.57 | 0.17 | 1.71 | 0.19 | 0.01 | 0.24 |
| VS-SC | 0.04 | 0.40 | 0.03 | 0.76 | 5.4 | 0.02 | 0.04 | -0.43 |
| DM-FP | 0.06 | 0.35 | -0.01 | 0.71 | 5.02 | 0.03 | 0.04 | -0.41 |
| AT-SM | 0.11 | -0.31 | -0.68 | 0.07 | 3.12 | 0.08 | 0.03 | 0.33 |

### 3.4. Temporal expression of brain CAPs: Occurrence rates

CAP occurrences measure how often each brain coactivation pattern appears during the fMRI scan. Higher occurrence rates indicate that a pattern occurs more frequently; lower rates suggest it appears less often. We examined baseline differences between groups (Section 3.4.1) and changes after 16 weeks of running therapy (Section 3.4.2).

#### 3.4.1. Baseline comparisons between controls and individuals with affective disorders

At baseline, we compared the occurrence rates of brain CAPs between healthy controls (n = 66) and individuals with affective disorders (n = 50). All five CAPs showed numerical differences between groups, but most yielded minor, non-significant effects (Figure 3B, Table 2, Table S13). The Visual-Sensorimotor-Subcortical CAP **(VS-SC_CAP_)** showed the most robust group difference, with reduced occurrences in the AFF group compared to controls (t-test: P= 0.04, d= 0.40, 95% CI [0.03, 0.76]; ANCOVA controlling for age and sex: F= 5.4, P= 0.02, η² = 0.04, β = -0.43). This medium-sized effect was the only finding with a 95% confidence interval that did not cross zero and remained significant after covariate adjustment. A comprehensive report of these baseline case-control analyses is presented in Table 2.

#### 3.4.2. Changes in brain CAPs and clinical symptoms after 16 weeks of running therapy

##### 3.4.2.1. Changes in Brain CAPs Induced by Treatment

Two CAPs showed significant VISIT × TREATMENT interactions: DM_CAP_ (F(1,41. 7) = 8.18, p = 0.01, partial η² = 0.13, Cohen’s d = –1.47. Table S15, S18) and VS-SC_CAP_ (F(1,41.6) = 8.10, p = 0.01, partial η² = 0.13, d = 1.47; Table S15, S18).

These effect sizes are large by conventional benchmarks and 1.75 times the minimum effect our sample could detect with 80% power (MDE(*d*) = 0.84; Table S16). Bootstrapped 95% CIs (5,000 iterations) excluded zero for both CAPs (DM: –2.15 to –0.29; VS-SC: 0.28 to 2.15; Table S15), confirming that even the lower confidence bounds constitute medium-to-large effects. Within-group changes were significant only after running therapy (DM: d = 0.88; VS-SC: d = –0.85; both p < 0.001) and negligible in the SSRI group (|d| ≤ 0.61, p ≥ 0.18; Table S17, Figure 5A), indicating treatment-specific reorganization rather than general time effects. Three additional CAPs displayed trend-level interactions (p = 0.06–0.22) with achieved power ≤ 37% (Table S16); these remain inconclusive.

##### 3.4.2.2. Brain CAPs-Symptom Associations at Baseline and Post-Treatment

We examined non-parametric correlations between the two treatment-responsive CAPs (DM_CAP_ and VS-SC_CAP_) and clinical symptom scores at baseline (T0) and post-treatment (T1) (Figure 5B). CAP-symptom correlations remained predominantly weak and non-significant at both timepoints. At baseline (T0), correlations ranged from |RHO|=0.01 to 0.24 (all P_FDR_>0.05). At post-treatment (T1), correlations ranged from |RHO|=0.03 to 0.43 (all P_FDR_>0.05), with the strongest association observed for DM and depression (ρ=−0.43). Although some correlation magnitudes increased from T0 to T1, none reached statistical significance after controlling for multiple comparisons.

###### Longitudinal Changes in CAP-Symptom Coupling

Next, we probed for changes in brain-symptom correlations from baseline to post-treatment. Correlations between the Default Mode CAP (DM_CAP_) and both symptom measures showed significant changes (DM_CAP_ × IDS: ΔRHO = -0.48, Z= -2.00, P= 0.05; DM_CAP_ × BAI: ΔRHO= -0.48, Z= -2.05, P= 0.04. Figure 5B), indicating moderate effects, confirmed by permutation testing (P_PERM_ = 0.06; 0.07. Complete results of changes in brain CAPs-symptom correlations between T0 and T1 are detailed in Table S19.

###### Brain Network Reorganization Does Not Predict Individual Symptom Improvement

These longitudinal changes in brain-behavior correlations prompted us to assess whether the degree of brain reorganization expressed by the CAPs occurrences was associated with symptom improvement (Figure S4). Among the two treatment-responsive CAPs, the Default Mode CAP (DM_CAP_) showed the largest, though non-significant, associations with clinical outcomes, but these correlations did not reach statistical significance (ΔDM_CAP_ × ΔIDS: RHO = -0.29, P = 0.17) and anxiety symptoms (ΔDM_CAP_ × ΔBAI: RHO = -0.31, P = 0.12. Figure S4).

## 4. DISCUSSION

We conducted a data-driven analysis (i.e., co-activation patterns (CAPs)) to characterize transient brain states during resting-state fMRI and their modulation by running therapy in patients with affective disorders. CAP analysis identified five distinct spatiotemporal configurations: AT-SM_CAP_ (salience-sensorimotor integration), DM_CAP_ (default mode network), VS-SC_CAP_ (frontoparietal control state, and a sensorimotor-visual pattern). While some CAPs overlapped with canonical intrinsic networks (e.g., DM_CAP_ with the default mode network), others demonstrated distributed architectures spanning multiple traditional static network boundaries (e.g., AT-SM_CAP_ combining salience, sensorimotor, and frontoparietal regions; VS-SC_CAP_ integrating visual cortex with somatomotor and subcortical structures. Critically, CAP metrics quantified temporal dynamics (occurrence rates) that fluctuated systematically with treatment, revealing network reconfigurations not captured by static connectivity approaches. This framework offers two key advantages: (1) it identifies functionally relevant brain states extending beyond canonical network parcellations, and (2) it captures transient fluctuations in brain network engagement that may reflect state-dependent changes in cognitive-affective processing relevant to treatment mechanisms in affective disorders.

### 4.1. Baseline Brain State Changes in Affective Disorders

At baseline, individuals with affective disorders exhibited fewer VS-SC_CAP_ occurrences compared to controls. This reinforces previous findings of reduced visual-somatosensory functional connectivity in depression (Guo et al., 2018). Similarly, activation in key brain regions involved in VS-SC_CAP_, such as the bilateral visual cortex (Moon et al., 2016) and bilateral sensorimotor cortex (Choi et al., 2012; Yang et al., 2017), has also been associated with anxiety disorders. However, evidence of visual-somatosensory functional connectivity related to anxiety remains limited. Moreover, concurrent disengagement of brain regions responsible for executive and self-referential functions (i.e., VIS-SC_CAP_ coactivation) has been observed in larger samples, in which these networks showed decreased within-network connectivity in 1300 Chinese individuals with MDD compared to 1128 controls from the REST-meta-MDD consortium (Yan et al., 2019). These results suggest transient disruptions in specific brain systems rather than constant, widespread alterations in the functional brain associated with affective disorders.

### 4.2. Running Therapy Induces Treatment-Specific Brain Network Reorganization

#### 4.2.1. DM_CAP_ and VS-SC_CAP_ Trajectories as Neural Substrates of Exercise-Induced Improvement

Running therapy induced significant reorganization of two brain networks: increased Default Mode CAP occurrence and decreased Visual-Somatosensory CAP occurrence (Figure 5A). These coordinated changes in self-referential and sensorimotor systems (Sheline et al., 2009; Zhang et al., 2024) were specific to running therapy, as the antidepressant group—serving as an active control with matched scanning protocols—showed minimal CAP changes despite comparable clinical improvement in the full MOTAR cohort (Verhoeven et al., 2023). Adequate statistical power for DM_CAP_ and VS-SC_CAP_ interaction effects (70% power, d=1.47) supports the reliability of these treatment-specific trajectories. Notably, these effects were undetectable using static connectivity methods in the same dataset (Vriend et al., 2025), indicating that therapeutic reorganization occurs in temporal network dynamics rather than spatial connectivity architecture. The coordinated DM_CAP_-VS-SC_CAP_ reconfiguration may reflect exercise-induced neuroplasticity, as running engages multiple neurobiological mechanisms, including neurotrophic signaling, neurogenesis, and anti-inflammatory effects (Cotman et al., 2007; Erickson et al., 2011). At the large-scale brain network level, exercise-induced changes in connectivity have been documented in the default mode and sensorimotor networks (Rajab et al., 2014; Schmitt et al., 2019, 2020). However, direct evidence linking molecular mechanisms to coordinated dynamic reorganization of these systems in affective disorders remains limited.

#### 4.2.2. Exercise-Induced Changes in Brain-Symptom Coupling

Although CAP-symptom correlations remained non-significant at both timepoints, stability analysis (Steiger’s Z-test) revealed that DMCAP-symptom relationships changed significantly between baseline and post-treatment: correlations with depression and anxiety shifted from weak positive/null at baseline to moderate negative post-treatment, indicating treatment-induced decoupling (Figure 5B, Table S19). In contrast, the visual-subcortical network (VS-SC_CAP_) exhibited correlation stability despite showing significant mean-level changes (Figure 5A). This suggests that VS-SCCAP changes may be dissociated from self-reported symptom measures or operate through mechanisms not captured by standard clinical questionnaires. These findings extend the established role of the default mode network as a diagnostic marker for affective disorders (Hamilton et al., 2015) by demonstrating that treatment-induced changes in DM spatiotemporal dynamics—specifically, its decoupling from symptoms and decreased occurrence following running therapy—constitute a potential brain biomarker for therapeutic response.

### 4.3. Implications for understanding exercise-induced brain plasticity

(1) Case-control comparisons suggest VS-SC_CAP_ may represent a trait-like vulnerability marker (reduced occurrences in patients vs. controls). On the other hand, the DM_CAP_ showed no baseline differences despite strong treatment responsiveness, demonstrating that spatiotemporal dynamics distinguish stable disease markers from treatment-sensitive brain states not captured by static connectivity. Replication in independent samples is needed to confirm these preliminary patterns. (2) Treatment-specific trajectories underscored that meaningful neurobiological change occurs in temporal variability of network engagement rather than spatial reconfiguration—a distinction captured only through dynamic connectivity and critical for differentiating exercise from pharmacotherapy mechanisms. (3) The DM_CAP_-symptoms associations, together with the absence of VS-SC_CAP_ correlations with self-reported measures observed in the RUN group, have two implications: First, they highlight DM_CAP_’s specificity as a biomarker of running treatment response. Second, they raise the question of whether substantial alterations in VS-SCCAP without symptom coupling reflect either (a) neuroplastic changes unrelated to core affective symptoms or (b) limitations of standard questionnaires in capturing exercise’s broader cognitive, somatic, and interoceptive benefits—a distinction that requires multimodal outcome assessment in future research.

## 5. LIMITATIONS AND FUTURE DIRECTIONS

The MRI subsample size limited our ability to examine disorder subtypes (e.g., immunometabolic depression, specific diagnoses) or conduct adequately powered within-group analyses in the antidepressant (AD) comparison group (N=9 post-treatment). Nonetheless, treatment-specific effects were robustly detected: VISIT × TREATMENT interactions for DMCAP and VS-SCCAP achieved 70% power with large effect sizes (d=1.47, partial η²=0.13; Tables S15-S16), and bootstrapped confidence intervals excluded zero (Table S15). The AD group served as an active control, demonstrating that running-induced neural reorganization reflects intervention-specific mechanisms rather than non-specific temporal effects (Nieuwenhuis et al., 2011).

Future research should employ larger samples to: (1) characterize neural reorganization patterns across affective disorder subtypes and clinical phenotypes; (2) examine dose-response relationships between exercise intensity/duration and brain network changes; and (3) integrate multilevel assessments linking molecular mechanisms (neurotrophic, inflammatory, dopaminergic signaling) to system-level brain dynamics to elucidate the pathways through which aerobic exercise ameliorates affective symptoms.

## 6. CONCLUSIONS

These findings indicate that running therapy alters brain network engagement patterns in individuals with affective disorders. By demonstrating that clinically effective interventions produce distinct dynamic reorganization signatures detectable only through time-resolved connectivity, this work positions temporal network dynamics as a framework for understanding the neural basis of treatment response and differentiating neuroplastic pathways across intervention modalities in affective disorders.

## Data Availability

All data produced in the present study are available upon reasonable request to the Principal Investigator of the MOTAR study

