## Supplementary material for "Running therapy improves clinical symptoms and reorganizes dynamic brain networks in affective disorders"

| **Authors** | Julian Gaviria Lopez *^a.^ ORCID ID: 0000-0002-4266-1371  Guido Van Wingen^a,d^. ORCID ID: 0000-0003-3076-5891  Chris Vriend^a,b,c^. ORCID ID: 0000-0003-3111-1304  Laura K.M. Han^a,d^. ORCID ID: 0000-0001-9647-3723  Jennifer Labus^e^. ORCID: 0000-0002-6634-2551  Gitte M. Knudsen^f,g^ ORCID ID: 0000-0003-1508-6866  Brenda W.J.H. Penninx ^a,d^. ORCID ID: 0000-0001-7779-9672  ^a^ Psychiatry department, Amsterdam UMC Location Vrije Universiteit, Amsterdam, the Netherlands. ^B^Department of Anatomy & Neurosciences, Amsterdam UMC, Location Vrije Universiteit Amsterdam, ^c^Amsterdam Neuroscience, Brain Imaging, Amsterdam, the Netherlands. Anatomy and Neurosciences, De Boelelaan 1117, Amsterdam, The Netherlands. ^d^Amsterdam Neuroscience, Mood, Anxiety, Psychosis, Sleep & Stress Program, Amsterdam, the Netherlands, ^e^G. Oppenheimer Center for Neurobiology of Stress and Resilience, Vatche & Tamar Manoukian Division of Digestive Diseases, Department of Medicine, David Geffen School of Medicine, University of California, Los Angeles, California.^f^Neurobiology Research Unit, Copenhagen University Hospital Rigshospitalet, Copenhagen, Denmark. e. ^g^Department of Clinical Medicine, Faculty of Health and Medicine Sciences, University of Copenhagen, Copenhagen, Denmark. |
| --- | --- |
| ***Correspondence** | Julian Gaviria Lopez. |
| **Running title** | Brain dynamics underlying running therapy |
| **Keywords** | Running therapy; Brain networks; affective disorders; dynamic functional connectivity (dFC); depression; anxiety. |

**Supplementary methods**

### **1. FMRI Preprocessing**

**1.1. Structural MRI Processing**: Structural MRI images were skull-stripped and segmented to reconstruct the brain surfaces using FreeSurfer 7.1.1 (Fischl, 2012). In case of longitudinal data (in individuals with an affective disorder only) we ran FreeSurfer on a mean robust template of the structural MRI scans of all available time points; otherwise, we used the structural MRI at baseline.

**1.2. Functional MRI Preprocessing:** Resting-state fMRI images were preprocessed using fMRIPrep v20.2.1 (Esteban et al., 2019; see supplementary methods for the boilerplate). Briefly, fMRI images from each time point were skull-stripped, realigned, slice-time corrected and corrected for susceptibility-induced distortions using a 'fieldmap-less' approach. Noise regressors were extracted for further denoising the time-series. We simultaneously performed denoising and band-pass filtering ([0.009–0.08 Hz]) using the denoiser tool (github.com/arielletambini/denoiser). We applied 'ICAAROMA8Phys' denoising that has previously been shown to provide a good denoising strategy in benchmark tests (Parkes et al., 2018). It consists of removing eight physiological signals from the white matter and cerebrospinal fluid (as well as their derivatives and quadratic terms) and the automatically identified motion-related components by automatic removal of motion artifacts using independent component analysis (ICA-AROMA) (Pruim et al., 2015).

**1.3. Motion Assessment and Quality Control:** We additionally calculated the framewise displacement (FD) using the Euclidean norm of translational and rotational head motion derivatives and excluded all participants with a mean root mean squared FD > 0.5 mm or >20 volumes with >0.5 mm volume-to-volume displacement, indicative of excessive motion (Power et al., 2012, 2014). For CAP analysis specifically, we implemented frame-level quality control: timepoints exceeding an FD threshold of 0.5 mm were identified as high-motion frames and excluded from subsequent clustering analysis (Figure 1B, corrupted frames shown as black dots), while low-motion frames were retained (Figure 1B, retained frames shown as gray dots). This frame-level censoring approach preserves usable data while minimizing motion-related confounds in the identification of brain co-activation patterns. Image quality metrics (e.g., DVARS and temporal SNR) were calculated using MRIqc (Esteban et al., 2017) and compared between groups and timepoints to ensure no systematic quality differences.

**1.4. Surface-Based Time Series Extraction:** Prior to time series extraction, the denoised functional images were mapped to the cortical surface. This approach has several advantages over volume-based approaches including better adherence to cortical convolutions and better spatial localization (Coalson et al., 2018; Dickie et al., 2019). For this we used the ciftify tool that allows applying 'human connectome project-style' processing to legacy data (Dickie et al., 2019) and projection of the denoised functional volumes to the cortical surface and resampling to fsLR32K standard space. The resulting cortical surface images were merged with segmentation maps of the 14 subcortical areas (derived from FreeSurfer). See (Dickie et al., 2019) for more details. No spatial smoothing was applied. Time series were extracted from 400 cortical brain areas that were parcellated according to the Schaefer atlas (Schaefer et al., 2018), and the 14 subcortical areas (thalamus, caudate, putamen, pallidum, hippocampus, amygdala, nucleus accumbens; bilateral), resulting in 414 nodes per participant.

### **2. Coactivation patterns (CAPs) generation and validation**

**2.1. CAP generation pipeline**: Frame selection and concatenation. For each participant, we extracted voxel-wise BOLD signal values from all retained low-motion frames identified during preprocessing (see Supplementary methods. “1. FMRI Preprocessing” section). Frames from all participants were combined into a single group-level matrix [i.e., participants x timepoints × voxels]. This method of concatenation ensures that the identified CAPs reflect patterns common across individuals rather than unique configurations. K-Means Clustering Implementation. We applied k-means clustering to the concatenated frame matrix to identify whole-brain co-activation states across the entire sample. Clustering parameters included: Euclidean distance (L2 norm), k-means++ initialization for optimal centroid placement, 500 maximum iterations (convergence criterion: 1×10⁻⁶), and 100 repetitions with different random initializations to avoid local minima. Clustering solutions were computed for k = 2–17, as previous studies have shown reliable cluster stability at k < 10 (Gaviria et al., 2021; Rey et al., 2021). Spatiotemporal metrics. After clustering, fMRI frames assigned to the same cluster (i.e., co-activation pattern (CAP) were averaged to generate spatial maps representing each CAP's configuration. These maps were then normalized by within-cluster standard error across volumes to produce Z-statistic maps, which quantified the degree to which each voxel's activation significantly deviated from zero. Critically, this approach characterizes both spatial and temporal properties of dynamic brain networks. Spatial CAP maps depict which brain regions co-activate (or co-deactivate) together, while temporal metrics quantify when and how often these patterns emerge. We computed CAP "occurrences"—the percentage of fMRI time frames during which each brain state was expressed. For example, if CAPx was present in 600 of 2,000 total frames, its occurrence would be 30%. Higher occurrence rates indicate a network state appears more often during the fMRI scan; lower rates suggest less frequent engagement (see illustration of this metric in Figure 2A). This temporal variability metric was used to examine both baseline group differences and treatment-induced changes in the dynamic expression of brain CAPs.

**2.2. Multi-criteria validation for selecting the optimal number of clusters.** A critical methodological challenge in CAP analysis is determining the optimal number of clusters (k) that balances model complexity with biological interpretability (Lurie et al., 2020). We employed three complementary validation approaches to triangulate the optimal clustering solution (Figure 2): A. Clustering Stability Analysis (Figure 2C). For each candidate k (2–17), we performed 100 k-means repetitions with random initializations. Clustering stability was quantified using consensus clustering, which measures the mean proportion of repetitions in which frame pairs were consistently assigned to the same cluster (range: 0 = unstable, 1 = perfectly stable). Solutions with stability indices below 0.22 (dashed line in Figure 2C) were considered unstable and excluded from further analysis. This threshold was determined empirically as two standard deviations below the mean stability of solutions with k = 3–7, which showed consistently high reproducibility (stability > 0.30). B. Intercluster Consistency Analysis (Figure 2D). We quantified agreement between partitions across k solutions using normalized mutual information (NMI) to assess whether fundamental patterns from simpler solutions (k=3,4,5) persisted in finer partitions (k=7–17). For each pair of solutions, the NMI was calculated as 2·MI(K₁, K₂)/[H(K₁) + H (K₂)], where H denotes Shannon entropy. High NMI scores (>0.7) indicate strong correspondence between CAPs. For instance, frames retained in CAPA at k=x generally remain together in CAPB at k=y—showing that the structure of this brain CAP is maintained across different clustering solutions. C. Biological Plausibility Assessment (Figure 2D). For each k, we quantified spatial correspondence with established neuroanatomy by calculating Dice overlap coefficients between CAP spatial maps and the Harvard-Oxford cortical (48 regions) and subcortical (21 regions) structural atlases (Desikan et al., 2006; Makris et al., 2006). The median Dice coefficient (range: 0 = no overlap, 1 = perfect overlap) across all CAP-atlas region pairs served as the biological plausibility score, with higher values indicating better alignment with known neuroanatomy. Summary of Validation Evidence. The convergence of spatial similarity (>90% overlap), temporal similarity (r >0.7), clustering stability (consensus >0.22), and biological plausibility (Dice ~0.80) establishes that the five identified brain CAPs represent stable, biologically grounded, and generalizable brain states suitable for investigating neural alterations related to affective disorders and running therapy.

**2.3. Validation across independent clustering scenarios.** To ensure that identified CAPs represent generalizable brain states rather than sample-specific artifacts, we assessed the robustness of the k solution resulting from the multi-criteria validation (cf. section 2.5.2.) across three independent clustering scenarios (Figure S1A): Scenario 1 - All Samples (N=116): Combined clustering using all participants (CTR + MDD) to derive group-representative CAPs. Scenario 2 - HC Group Only (N=66): Clustering performed exclusively on healthy control data. Scenario 3 - MDD Group Only (N=50): Clustering performed exclusively on patient data. Note: Hereafter, "MDD" refers collectively to all patients with affective disorders (major depressive disorder, anxiety disorders, or comorbid presentations). Spatial Similarity Assessment (Figure S1B). We quantified spatial correspondence between CAPs identified in each scenario using Dice overlap coefficients and Jaccard indices. Both metrics demonstrated remarkably high overlap (>94% Dice, >88% Jaccard) across all scenario comparisons, indicating that the five CAPs represent fundamental brain states present in both healthy and clinical populations rather than being driven by either group's unique characteristics. Temporal Similarity Assessment (Figure S1C). Beyond spatial pattern matching, we assessed whether CAPs exhibited similar temporal dynamics across scenarios by calculating Pearson correlation coefficients (r) and concordance correlation coefficients (CCC) for CAP occurrence frequencies. Heatmaps display pairwise correlations between CAPs across scenarios, with strong diagonal correlations (r >0.7, dark magenta) confirming that each CAP from one scenario has a clear temporal correspondent in other scenarios. High concordance coefficients (CCC >0.7) and low bias factors indicate consistent temporal expression characteristics across samples.

### **5. Supplementary Figures**

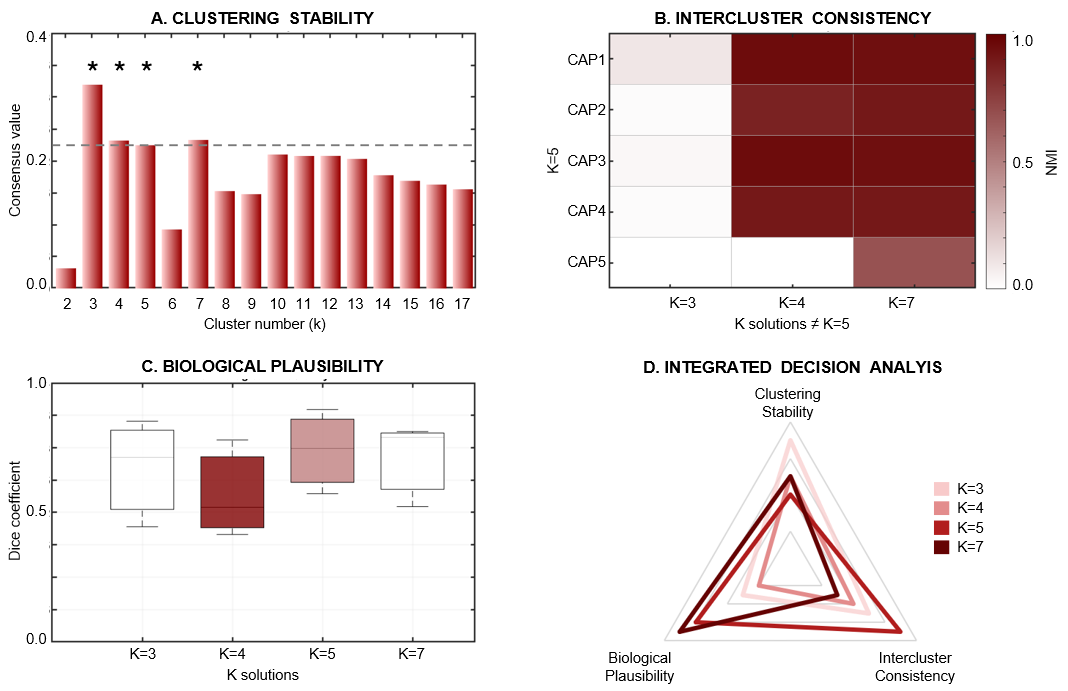

**5.1. Supplementary Figure S1. Multi-criteria validation of CAP clustering.** **A.** Clustering stability: Consensus stability index across cluster numbers (K=2-17). Asterisks indicate highest stability solutions (K=3, 4, 5, 7). **B.** Intercluster consistency: Normalized mutual information (NMI) quantifies spatial correspondence between CAPs across different K solutions. High consistency (dark red) between K=4-5, and K=5-7 indicates robust clustering at K=5. C. Biological plausibility: Dice overlap coefficients between CAP spatial maps and neuroanatomical atlas networks (Harvard-Oxford atlas). Higher values indicate better neuroanatomical alignment. D. Integrated decision analysis: Radar plot synthesizes three validation criteria (stability, consistency, plausibility) across K solutions. Convergent evidence supports K=5 as the optimal solution.

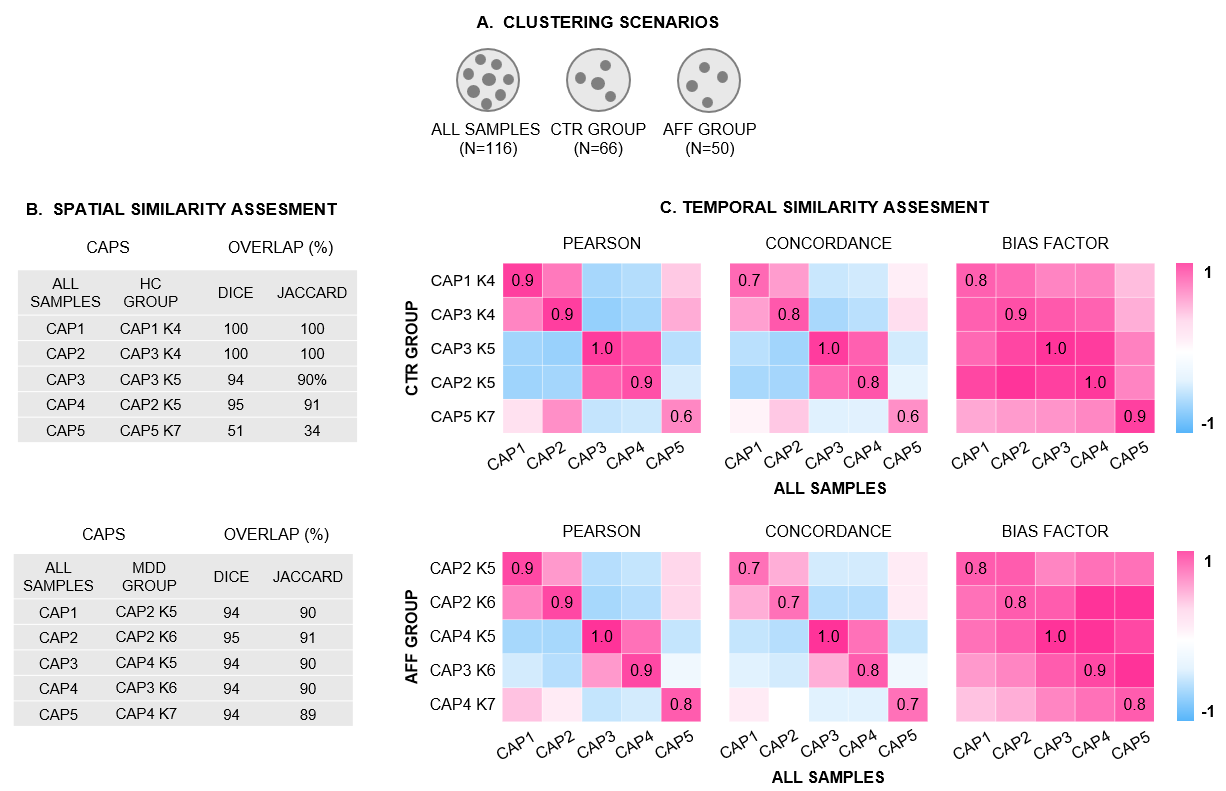

**5.2. Supplementary Figure S2**. **Co-Activation Pattern Validation Across Clustering Scenarios. A.** Clustering scenarios: CAPs were derived from three independent clustering approaches: all samples combined (N=116), healthy controls only (CTR, N=66), and participants with affective disorders (AFF, N=50). **B.** Spatial similarity assessment quantifies overlap between CAPs identified in all-samples versus group-specific clustering (z-score > |1.3|) using Dice and Jaccard coefficients. High overlap (>90%) indicates robust and reproducible spatial patterns across clustering scenarios. **C.** Temporal similarity assessment evaluates correspondence of CAP occurrences across clustering approaches via Pearson correlation and concordance correlation coefficients, including their bias factor respectively. Strong positive correlations (magenta, r >0.8) and high concordance demonstrate consistent temporal dynamics across scenarios, validating the robustness of identified CAPs independent of sample composition.

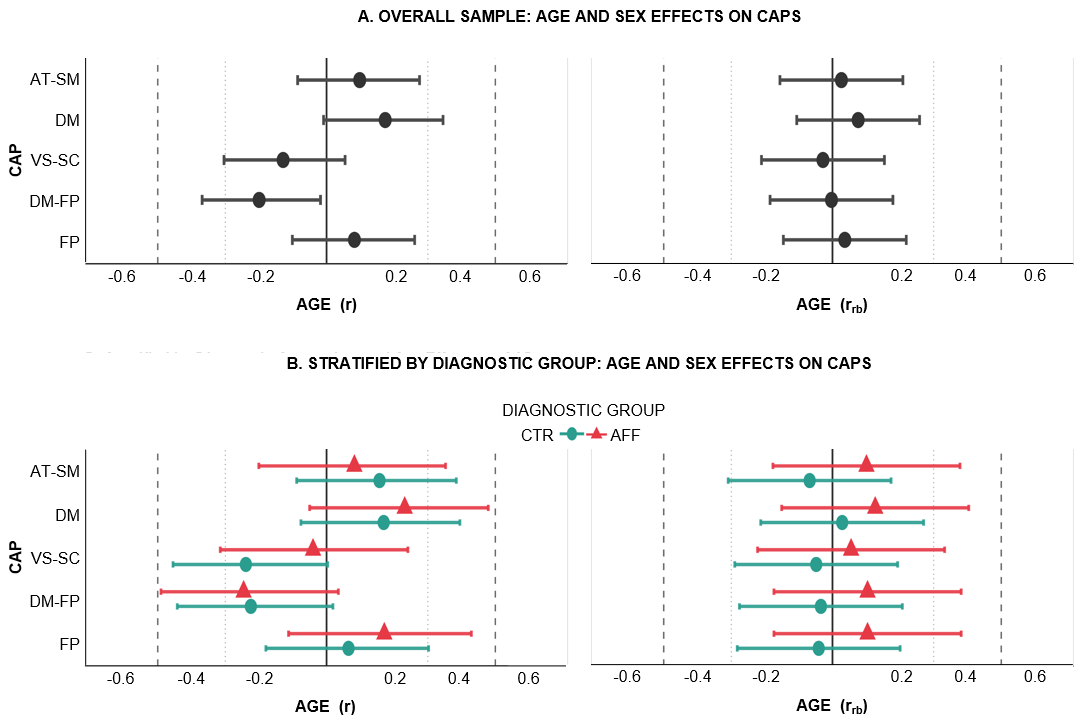

**5.3. Supplementary Figure S3. Demographic Associations with CAP Temporal Occurrence**. **A.** **Overall sample** (N=116: 66 controls (CTR group), 50 individual with affective disorders (AFF group)). Age: Pearson r (95% CI). Sex: rank-biserial r_rb_ (95% CI) from Mann-Whitney U test (58 males, 58 females). All effect sizes |r| or |r_rb_|< 0.2 (negligible-to-small), indicating minimal demographic associations. **B. Stratified by diagnostic group.** Controls: n=66 (36M/30F); AFF: N=50 (22M/28F). No significant effects in either group (all P> 0.20), consistent with minimal demographic confounding. Solid line = zero effect; dashed lines = large effect (|r|≥0.5); dotted lines = medium effect (|r|≥0.3). Error bars = 95% CI. *P<0.05; **P<0.01; ***P<0.001. Green circles = Controls; red triangles = AFF.

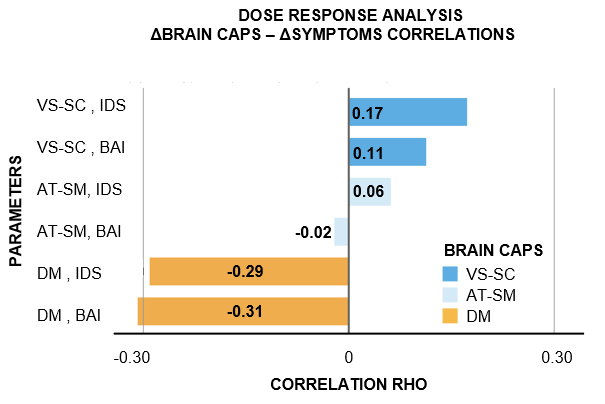

**5.4. Supplementary Figure S4**. **Dose-response coupling: correlations between brain network changes and symptom improvements.** We quantified the coupling between the magnitude of brain network reorganization and symptom relief to examine whether individual differences in CAP changes predicted symptom improvement following running therapy (N=26 paired observations). Change scores were calculated for each subject (ΔCAP = CAP_T1 - CAP_T0; ΔSymptom = Symptom_T1 - Symptom_T0) and correlated using Spearman's rank correlation coefficient. Among the three treatment-responsive CAPs, decreased occurrence rates of the default mode (DM) CAP showed the strongest correlations with symptom improvements: larger DM_CAP_ reductions predicted greater depression improvement (ΔDM × ΔIDS: RHO = -0.29, P = 0.17) and greater anxiety reduction (ΔDM × ΔBAI: RHO= -0.31, P= 0.12). In contrast, changes in attention-somatomotor (AT-SM_CAP_) and ventral salience-subcortical areas (VS-SC_CAP_) showed minimal coupling with symptom changes (all |ρ| < 0.20, all p > 0.45), suggesting dissociation between these networks' reorganization and therapeutic benefit.

### **6. Supplementary tables**

| **AT-SM_CAP_ COACTIVATION** | | | | | |
| --- | --- | --- | --- | --- | --- |
| **Region_AAL3** | **Hemisphere** | **Voxels** | **Percent Total** | **Yeo17_Network** | **Functional System** |
| SupraMarginal_R | Right | 1403 | 6.2 | SomMotB | Sensorimotor |
| Insula_R | Right | 1300 | 5.7 | SalVentAttnA | Salience |
| Insula_L | Left | 1194 | 5.3 | SalVentAttnA | Salience |
| Supp_Motor_Area_R | Right | 1084 | 4.8 | SomMotB | Sensorimotor |
| Rolandic_Oper_R | Right | 1037 | 4.6 | SomMotB | Sensorimotor |
| Temporal_Sup_L | Left | 1013 | 4.5 | TempPar | Temporoparietal |
| Supp_Motor_Area_L | Left | 938 | 4.1 | SomMotB | Sensorimotor |
| SupraMarginal_L | Left | 924 | 4.1 | SomMotB | Sensorimotor |
| Cingulate_Mid_R | Right | 911 | 4 | SalVentAttnB | Salience |
| Precentral_R | Right | 734 | 3.2 | SomMotA | Sensorimotor |
| Rolandic_Oper_L | Left | 714 | 3.1 | SomMotB | Sensorimotor |
| Frontal_Inf_Oper_R | Right | 684 | 3 | ContC | Control |
| Temporal_Sup_R | Right | 680 | 3 | TempPar | Temporoparietal |
| Postcentral_R | Right | 666 | 2.9 | SomMotA | Sensorimotor |
| Cingulate_Mid_L | Left | 650 | 2.9 | SalVentAttnB | Salience |
| Frontal_Sup_2_R | Right | 603 | 2.7 | ContA | Control |
| Postcentral_L | Left | 561 | 2.5 | SomMotA | Sensorimotor |
| Frontal_Mid_2_R | Right | 468 | 2.1 | ContA | Control |
| Frontal_Mid_2_L | Left | 467 | 2.1 | ContA | Control |
| Frontal_Inf_Oper_L | Left | 401 | 1.8 | ContC | Control |
| Parietal_Inf_L | Left | 375 | 1.7 | ContB | Control |
| Frontal_Sup_2_L | Left | 307 | 1.4 | ContA | Control |
| Parietal_Sup_L | Left | 255 | 1.1 | DorsAttnB | Attention |
| Parietal_Inf_R | Right | 245 | 1.1 | ContB | Control |
| Parietal_Sup_R | Right | 232 | 1 | DorsAttnB | Attention |
| Precentral_L | Left | 192 | 0.8 | SomMotA | Sensorimotor |
| Precuneus_L | Left | 187 | 0.8 | DefaultC | DefaultMode |
| Precuneus_R | Right | 182 | 0.8 | DefaultC | DefaultMode |
| Temporal_Pole_Sup_R | Right | 178 | 0.8 | LimbicA | Limbic |
| Frontal_Mid_2_R | Right | 174 | 0.8 | ContA | Control |
| Precentral_L | Left | 169 | 0.7 | SomMotA | Sensorimotor |
| Frontal_Inf_Tri_R | Right | 155 | 0.7 | ContA | Control |
| ACC_sup_L | Left | 154 | 0.7 | SalVentAttnA | Salience |
| Temporal_Pole_Sup_L | Left | 146 | 0.6 | LimbicA | Limbic |
| Temporal_Mid_L | Left | 135 | 0.6 | DefaultA | DefaultMode |
| Cingulate_Mid_L | Left | 127 | 0.6 | SalVentAttnB | Salience |
| Heschl_L | Left | 122 | 0.5 | Auditory | Auditory |
| Putamen_R | Right | 125 | 0.6 | Subcortical | Subcortical |
| Cerebellum_8_L | Left | 111 | 0.5 | Subcortical | Cerebellum |
| Heschl_R | Right | 98 | 0.4 | Auditory | Auditory |
| Frontal_Mid_2_R | Right | 93 | 0.4 | ContA | Control |
| Frontal_Sup_2_R | Right | 89 | 0.4 | ContA | Control |
| Frontal_Inf_Tri_L | Left | 88 | 0.4 | ContA | Control |
| Paracentral_Lobule_L | Left | 75 | 0.3 | SomMotA | Sensorimotor |
| ACC_sup_R | Right | 70 | 0.3 | SalVentAttnA | Salience |
| Paracentral_Lobule_R | Right | 67 | 0.3 | SomMotA | Sensorimotor |
| Frontal_Inf_Orb_2_R | Right | 60 | 0.3 | LimbicB | Limbic |
| Frontal_Inf_Orb_2_L | Left | 41 | 0.2 | LimbicB | Limbic |
| Temporal_Mid_R | Right | 40 | 0.2 | DefaultA | DefaultMode |
| Frontal_Sup_Medial_L | Left | 18 | 0.1 | DefaultB | DefaultMode |
| Putamen_R | Left | 14 | 0.1 | Subcortical | Subcortical |

**6.1. Supplementary Table S1. CAP1 (attention-sensorimotor) coactivation pattern: Regional composition and functional network assignment.** Anatomical regions comprising AT-SM_CAP_ co-activation pattern (Z > 1.3, cluster ≥ 30 voxels; 22,676 total voxels). Region_AAL3. Automated Anatomical Labeling Atlas v3.1 nomenclature (L, left; R, right). Hemisphere: Lateralization of anatomical region. Voxels: Number of 2mm³ MNI voxels per region. Percent Total: Proportion of CAP1 total voxels (e.g., SupraMarginal_R: 1,403 voxels = 6.2%). Yeo17_Network: Assignment to Yeo's 17-network parcellation (Schaefer et al., 2018) – SomMotA/B (Somatomotor), SalVentAttnA/B (Salience/Ventral Attention), TempPar (Temporoparietal), ContA/B/C (Control), DorsAttnB (Dorsal Attention), DefaultA/B/C (Default Mode), LimbicA/B (Limbic), Auditory, Subcortical (Basal Ganglia, Cerebellum). Functional System: Higher-level aggregation into cognitive systems (Sensorimotor, Salience, Temporoparietal, Control, Attention, DefaultMode, Limbic, Auditory, Subcortical, Cerebellum).

| **AT-SM_CAP_ (CODEACTIVATION)** | | | | | |
| --- | --- | --- | --- | --- | --- |
| **Region_AAL3** | **Hemisphere** | **Voxels** | **Percent Total** | **Yeo17_Network** | **Functional System** |
| Precuneus_L | Left | 988 | 14.7 | DefaultC | DefaultMode |
| Angular_L | Left | 805 | 12 | DefaultC | DefaultMode |
| Frontal_Sup_2_L | Left | 654 | 9.7 | ContA | Control |
| Angular_R | Right | 706 | 10.5 | DefaultC | DefaultMode |
| Precuneus_R | Right | 615 | 9.1 | DefaultC | DefaultMode |
| Frontal_Sup_2_R | Right | 226 | 3.4 | ContA | Control |
| Temporal_Mid_L | Left | 268 | 4 | DefaultA | DefaultMode |
| Frontal_Med_Orb_L | Left | 195 | 2.9 | DefaultB | DefaultMode |
| Cingulate_Post_L | Left | 185 | 2.8 | DefaultC | DefaultMode |
| Cingulate_Mid_L | Left | 196 | 2.9 | DefaultC | DefaultMode |
| Cingulate_Mid_R | Right | 152 | 2.3 | DefaultC | DefaultMode |
| Temporal_Mid_R | Right | 124 | 1.8 | DefaultA | DefaultMode |
| Cerebellum_Crus1_R | Right | 123 | 1.8 | Subcortical | Cerebellum |
| Frontal_Med_Orb_R | Right | 104 | 1.5 | DefaultB | DefaultMode |
| Parietal_Inf_L | Left | 98 | 1.5 | DefaultC | DefaultMode |
| Calcarine_L | Left | 98 | 1.5 | VisCent | Visual |
| Cerebellum_Crus1_L | Left | 40 | 0.6 | Subcortical | Cerebellum |
| Cerebellum_Crus2_R | Right | 34 | 0.5 | Subcortical | Cerebellum |
| Cerebellum_9_R | Right | 32 | 0.5 | Subcortical | Cerebellum |
| Cuneus_L | Left | 147 | 2.2 | VisPeri | Visual |
| Calcarine_R | Right | 51 | 0.8 | VisCent | Visual |
| Cingulate_Post_R | Right | 65 | 1 | DefaultC | DefaultMode |
| Rectus_L | Left | 107 | 1.6 | DefaultB | DefaultMode |
| Rectus_R | Right | 81 | 1.2 | DefaultB | DefaultMode |
| Frontal_Sup_Medial_L | Left | 90 | 1.3 | DefaultB | DefaultMode |
| Frontal_Sup_Medial_R | Right | 26 | 0.4 | DefaultB | DefaultMode |
| Frontal_Mid_2_L | Left | 350 | 5.2 | ContA | Control |
| Occipital_Mid_L | Left | 98 | 1.5 | VisPeri | Visual |
| Parietal_Inf_R | Right | 15 | 0.2 | DefaultC | DefaultMode |

**6.2. Supplementary Table S2. CAP1 (attention-sensorimotor) co-deactivation pattern**: **Regional suppression during attention-sensorimotor engagement.** Anatomical regions exhibiting negative BOLD signal (deactivation/suppression) during AT-SM_CAP_ occurrences (Z < -1.3, cluster ≥ 30 voxels; 6,717 total voxels). Region_AAL3: Automated Anatomical Labeling Atlas v3.1 nomenclature (L, left; R, right). Hemisphere: Lateralization of anatomical region. Voxels: Number of 2mm³ MNI voxels per region showing coordinated deactivation. Percent Total: Proportion of CAP1 total deactivated voxels (e.g., Precuneus_L: 988 voxels = 14.7%). Yeo17_Network: Assignment to Yeo's 17-network parcellation (Schaefer et al., 2018) – DefaultA/B/C (Default Mode A/B/C), ContA (Control A), VisCent/VisPeri (Visual Central/Peripheral), Subcortical (Cerebellum). Functional System: Higher-level aggregation into cognitive systems (DefaultMode, Control, Visual, Cerebellum). Network Profile: AT-SM_CAP_ co-deactivation predominantly engages Default Mode Network (81.7%), including posterior DMN hubs (Precuneus, Angular gyrus, Posterior Cingulate: 66.3%) and anterior DMN (medial Prefrontal, Orbitofrontal: 9.9%), alongside Frontoparietal Control regions (13.1%), Visual cortex (4.4%), and Cerebellum (3.5%). This reciprocal pattern—activation of sensorimotor-salience systems (Table S1) coupled with deactivation of internally-directed default mode networks—characterizes a brain state prioritizing external sensory processing and bodily awareness over internal mentation and self-referential thought.

| **DM_CAP_ (COACTIVATION)** | | | | | |
| --- | --- | --- | --- | --- | --- |
| **Region_AAL3** | **Hemisphere** | **Voxels** | **Percent Total** | **Yeo17_Network** | **Functional System** |
| Frontal_Sup_Medial_L | Left | 1349 | 13.1 | DefaultB | DefaultMode |
| Precuneus_L | Left | 929 | 9 | DefaultC | DefaultMode |
| Frontal_Sup_Medial_R | Right | 853 | 8.3 | DefaultB | DefaultMode |
| Angular_L | Left | 794 | 7.7 | DefaultC | DefaultMode |
| Temporal_Mid_L | Left | 747 | 7.2 | DefaultA | DefaultMode |
| Precuneus_R | Right | 658 | 6.4 | DefaultC | DefaultMode |
| Angular_R | Right | 631 | 6.1 | DefaultC | DefaultMode |
| Temporal_Mid_R | Right | 468 | 4.5 | DefaultA | DefaultMode |
| Frontal_Med_Orb_L | Left | 388 | 3.8 | DefaultB | DefaultMode |
| Frontal_Sup_2_L | Left | 335 | 3.2 | ContA | Control |
| Frontal_Sup_2_R | Right | 212 | 2.1 | ContA | Control |
| Rectus_L | Left | 261 | 2.5 | DefaultB | DefaultMode |
| Frontal_Med_Orb_R | Right | 257 | 2.5 | DefaultB | DefaultMode |
| Cingulate_Post_L | Left | 186 | 1.8 | DefaultC | DefaultMode |
| Temporal_Pole_Mid_R | Right | 183 | 1.8 | LimbicA | Limbic |
| Cerebellum_Crus1_R | Right | 179 | 1.7 | Subcortical | Cerebellum |
| Rectus_R | Right | 173 | 1.7 | DefaultB | DefaultMode |
| Cingulate_Mid_L | Left | 136 | 1.3 | DefaultC | DefaultMode |
| Cingulate_Mid_R | Right | 113 | 1.1 | DefaultC | DefaultMode |
| Cerebellum_Crus1_L | Left | 112 | 1.1 | Subcortical | Cerebellum |
| Cuneus_L | Left | 114 | 1.1 | VisPeri | Visual |
| Temporal_Pole_Mid_L | Left | 94 | 0.9 | LimbicA | Limbic |
| ACC_pre_L | Left | 90 | 0.9 | DefaultB | DefaultMode |
| Cingulate_Post_R | Right | 79 | 0.8 | DefaultC | DefaultMode |
| ACC_pre_R | Right | 72 | 0.7 | DefaultB | DefaultMode |
| Occipital_Mid_L | Left | 53 | 0.5 | VisPeri | Visual |
| Temporal_Mid_L | Left | 48 | 0.5 | DefaultA | DefaultMode |
| Cerebellum_9_R | Right | 42 | 0.4 | Subcortical | Cerebellum |
| Cerebellum_Crus2_R | Right | 42 | 0.4 | Subcortical | Cerebellum |
| Cerebellum_Crus2_L | Left | 44 | 0.4 | Subcortical | Cerebellum |
| Temporal_Inf_R | Right | 39 | 0.4 | DefaultA | DefaultMode |
| Temporal_Mid_R | Right | 32 | 0.3 | DefaultA | DefaultMode |
| Temporal_Sup_R | Right | 29 | 0.3 | TempPar | Temporoparietal |
| Calcarine_L | Left | 25 | 0.2 | VisCent | Visual |
| Temporal_Inf_L | Left | 21 | 0.2 | DefaultA | DefaultMode |
| Parietal_Inf_L | Left | 18 | 0.2 | DefaultC | DefaultMode |
| Parietal_Inf_R | Right | 17 | 0.2 | DefaultC | DefaultMode |
| Occipital_Mid_R | Right | 15 | 0.1 | VisPeri | Visual |
| Calcarine_R | Right | 14 | 0.1 | VisCent | Visual |
| Temporal_Pole_Sup_R | Right | 14 | 0.1 | LimbicA | Limbic |
| Cuneus_R | Right | 9 | 0.1 | VisPeri | Visual |
| Temporal_Sup_L | Left | 5 | 0 | TempPar | Temporoparietal |
| SupraMarginal_L | Left | 4 | 0 | SomMotB | Sensorimotor |

**6.3. Supplementary Table S3. CAP2 (default mode) co-activation pattern:** Regional Composition and Functional Network Assignment. Anatomical regions comprising DM_CAP_ co-activation pattern (Z > 1.3, cluster ≥ 30 voxels; 10,319 total voxels). Region_AAL3: Automated Anatomical Labeling Atlas v3.1 nomenclature (L, left; R, right). Voxels: Number of 2mm³ MNI voxels per region. Percent Total: Proportion of CAP2 total voxels (e.g., Frontal_Sup_Medial_L: 1,349 voxels = 13.1%). Yeo17_Network: Assignment to Yeo's 17-network parcellation (Schaefer et al., 2018) – DefaultA/B/C (Default Mode), ContA (Control), LimbicA/B (Limbic), VisCent/VisPeri (Visual Central/Peripheral), Subcortical (Cerebellum), TempPar (Temporoparietal). Functional System: Higher-level aggregation into cognitive systems (DefaultMode, Control, Limbic, Visual, Cerebellum, Temporoparietal). Network Profile: DM_CAP_ integrates Default Mode Network subsystems including Anterior DM (medial Prefrontal, Orbitofrontal: 35.4%), Posterior DMN (Precuneus, Angular gyrus, Posterior Cingulate: 43.7%), Temporal DM (Middle Temporal Gyrus: 16.6%), Control (5.3%), Cerebellum (4.1%), Limbic (2.8%), and Visual (2.0%) regions, characterizing a transient state supporting internally-directed cognition. This multi-system configuration supports self-referential thought, episodic memory retrieval, future simulation, and social cognition—processes predominant during wakeful rest and spontaneous mind-wandering.

| **DM_CAP_ (CO-DEACTIVATION)** | | | | | |
| --- | --- | --- | --- | --- | --- |
| **Region_AAL3** | **Hemisphere** | **Voxels** | **Percent Total** | **Yeo17_Network** | **Functional System** |
| Parietal_Inf_L | Left | 1220 | 8.4 | ContB | Control |
| Insula_R | Right | 609 | 4.2 | SalVentAttnA | Salience |
| Frontal_Mid_2_R | Right | 988 | 6.8 | ContA | Control |
| Frontal_Inf_Oper_R | Right | 648 | 4.5 | ContC | Control |
| Insula_L | Left | 596 | 4.1 | SalVentAttnA | Salience |
| Parietal_Sup_R | Right | 622 | 4.3 | DorsAttnB | Attention |
| SupraMarginal_R | Right | 1044 | 7.2 | SomMotB | Sensorimotor |
| Frontal_Mid_2_L | Left | 476 | 3.3 | ContA | Control |
| SupraMarginal_L | Left | 467 | 3.2 | SomMotB | Sensorimotor |
| Frontal_Sup_2_R | Right | 399 | 2.8 | ContA | Control |
| Parietal_Sup_L | Left | 381 | 2.6 | DorsAttnB | Attention |
| Frontal_Inf_Tri_R | Right | 212 | 1.5 | ContA | Control |
| Frontal_Inf_Oper_L | Left | 368 | 2.5 | ContC | Control |
| Frontal_Inf_Tri_L | Left | 392 | 2.7 | ContA | Control |
| Supp_Motor_Area_R | Right | 293 | 2 | SomMotB | Sensorimotor |
| Supp_Motor_Area_L | Left | 251 | 1.7 | SomMotB | Sensorimotor |
| Frontal_Sup_2_L | Left | 187 | 1.3 | ContA | Control |
| Temporal_Mid_L | Left | 169 | 1.2 | DefaultA | DefaultMode |
| Cingulate_Mid_R | Right | 160 | 1.1 | SalVentAttnB | Salience |
| Postcentral_R | Right | 161 | 1.1 | SomMotA | Sensorimotor |
| Temporal_Inf_L | Left | 125 | 0.9 | DefaultA | DefaultMode |
| Temporal_Inf_R | Right | 111 | 0.8 | DefaultA | DefaultMode |
| Rolandic_Oper_R | Right | 143 | 1 | SomMotB | Sensorimotor |
| Postcentral_L | Left | 107 | 0.7 | SomMotA | Sensorimotor |
| Precuneus_L | Left | 104 | 0.7 | DefaultC | DefaultMode |
| Precentral_R | Right | 53 | 0.4 | SomMotA | Sensorimotor |
| Precentral_L | Left | 83 | 0.6 | SomMotA | Sensorimotor |
| Cingulate_Mid_L | Left | 64 | 0.4 | SalVentAttnB | Salience |
| Rolandic_Oper_L | Left | 63 | 0.4 | SomMotB | Sensorimotor |
| Temporal_Sup_L | Left | 31 | 0.2 | TempPar | Temporoparietal |
| Temporal_Pole_Sup_R | Right | 58 | 0.4 | LimbicA | Limbic |
| Temporal_Pole_Sup_L | Left | 64 | 0.4 | LimbicA | Limbic |
| Angular_R | Right | 73 | 0.5 | DefaultC | DefaultMode |
| Precuneus_R | Right | 53 | 0.4 | DefaultC | DefaultMode |
| Cerebellum_8_L | Left | 53 | 0.4 | Subcortical | Cerebellum |
| Cerebellum_8_R | Right | 40 | 0.3 | Subcortical | Cerebellum |
| Cerebellum_7b_L | Left | 26 | 0.2 | Subcortical | Cerebellum |
| Occipital_Mid_L | Left | 47 | 0.3 | VisPeri | Visual |
| Occipital_Sup_R | Right | 47 | 0.3 | VisPeri | Visual |
| Occipital_Inf_L | Left | 13 | 0.1 | VisPeri | Visual |
| Temporal_Mid_R | Right | 18 | 0.1 | DefaultA | DefaultMode |
| Frontal_Inf_Orb_2_R | Right | 22 | 0.2 | LimbicB | Limbic |

**6.4. Supplementary Table S4. CAP2 (default mode) co-deactivation pattern**: **Regional suppression during default mode engagement.** Anatomical regions exhibiting negative BOLD signal (deactivation/suppression) during DM_CAP_ occurrences (Z < -1.3, cluster ≥ 30 voxels; 14,508 total voxels). Region_AAL3: Automated Anatomical Labeling Atlas v3.1 nomenclature (L, left; R, right). Hemisphere: Lateralization of anatomical region. Voxels: Number of 2mm³ MNI voxels per region showing coordinated deactivation. Percent Total: Proportion of CAP2 total deactivated voxels (e.g., Parietal_Inf_L: 1,220 voxels = 8.4%). Yeo17 Network: Assignment to Yeo's 17-network parcellation (Schaefer et al., 2018) – ContA/B/C (Control), SalVentAttnA/B (Salience/Ventral Attention), SomMotA/B (Somatomotor), DorsAttnB (Dorsal Attention), DefaultA/C (Default Mode), LimbicA/B (Limbic), TempPar (Temporoparietal), Subcortical (Cerebellum), VisPeri (Visual Peripheral). Functional System: Higher-level aggregation into cognitive systems (Control, Sensorimotor, Salience, Attention, DefaultMode, Limbic, Temporoparietal, Cerebellum, Visual). Network Profile: DM_CAP_ co-deactivation predominantly engages Frontoparietal Control (35.7%), Sensorimotor (18.0%), Salience (9.8%), and Dorsal Attention (6.9%) networks, characterizing a reciprocal suppression pattern during internally-directed cognition.

| **VS-SC_CAP_ COACTIVATION** | | | | | |
| --- | --- | --- | --- | --- | --- |
| **Region_AAL3** | **Hemisphere** | **Voxels** | **Percent Total** | **Yeo17_Network** | **Functional System** |
| Occipital_Mid_L | Left | 2318 | 7.2 | VisPeri | Visual |
| Calcarine_L | Left | 1961 | 6.1 | VisCent | Visual |
| Lingual_R | Right | 1941 | 6 | VisCent | Visual |
| Postcentral_R | Right | 1889 | 5.8 | SomMotA | Sensorimotor |
| Lingual_L | Left | 1824 | 5.6 | VisCent | Visual |
| Postcentral_L | Left | 1810 | 5.6 | SomMotA | Sensorimotor |
| Calcarine_R | Right | 1598 | 4.9 | VisCent | Visual |
| Occipital_Mid_R | Right | 1559 | 4.8 | VisPeri | Visual |
| Cuneus_R | Right | 1287 | 4 | VisPeri | Visual |
| Cuneus_L | Left | 1249 | 3.9 | VisPeri | Visual |
| Fusiform_R | Right | 1210 | 3.7 | VisPeri | Visual |
| Occipital_Sup_L | Left | 1153 | 3.6 | VisPeri | Visual |
| Precentral_R | Right | 1091 | 3.4 | SomMotA | Sensorimotor |
| Occipital_Sup_R | Right | 1081 | 3.3 | VisPeri | Visual |
| Fusiform_L | Left | 989 | 3.1 | VisPeri | Visual |
| Occipital_Inf_R | Right | 808 | 2.5 | VisPeri | Visual |
| Occipital_Inf_L | Left | 770 | 2.4 | VisPeri | Visual |
| Temporal_Mid_R | Right | 662 | 2 | DefaultA | DefaultMode |
| Parietal_Sup_R | Right | 564 | 1.7 | DorsAttnB | Attention |
| Precentral_L | Left | 559 | 1.7 | SomMotA | Sensorimotor |
| Parietal_Sup_L | Left | 460 | 1.4 | DorsAttnB | Attention |
| Paracentral_Lobule_L | Left | 347 | 1.1 | SomMotA | Sensorimotor |
| Temporal_Inf_R | Right | 324 | 1 | DefaultA | DefaultMode |
| Paracentral_Lobule_R | Right | 314 | 1 | SomMotA | Sensorimotor |
| Temporal_Mid_L | Left | 262 | 0.8 | DefaultA | DefaultMode |
| Cerebellum_6_L | Left | 191 | 0.6 | Subcortical | Cerebellum |
| Precuneus_L | Left | 183 | 0.6 | DefaultC | DefaultMode |
| Cerebellum_6_R | Right | 157 | 0.5 | Subcortical | Cerebellum |
| Precuneus_R | Right | 159 | 0.5 | DefaultC | DefaultMode |
| Supp_Motor_Area_R | Right | 152 | 0.5 | SomMotB | Sensorimotor |
| Parietal_Inf_L | Left | 111 | 0.3 | ContB | Control |
| Cerebellum_4_5_R | Right | 104 | 0.3 | Subcortical | Cerebellum |
| ParaHippocampal_R | Right | 103 | 0.3 | LimbicA | Limbic |
| Cerebellum_4_5_L | Left | 87 | 0.3 | Subcortical | Cerebellum |
| Parietal_Sup_R | Right | 82 | 0.3 | DorsAttnB | Attention |
| Precuneus_L | Left | 77 | 0.2 | DefaultC | DefaultMode |
| Precuneus_R | Right | 75 | 0.2 | DefaultC | DefaultMode |
| Rolandic_Oper_L | Left | 55 | 0.2 | SomMotB | Sensorimotor |
| Supp_Motor_Area_L | Left | 51 | 0.2 | SomMotB | Sensorimotor |
| Parietal_Sup_L | Left | 45 | 0.1 | DorsAttnB | Attention |
| Vermis_4_5 | Midline | 33 | 0.1 | Subcortical | Cerebellum |
| Temporal_Inf_L | Left | 32 | 0.1 | DefaultA | DefaultMode |
| ParaHippocampal_L | Left | 24 | 0.1 | LimbicA | Limbic |
| Frontal_Mid_2_R | Right | 20 | 0.1 | ContA | Control |
| SupraMarginal_L | Left | 15 | 0 | SomMotB | Sensorimotor |
| Frontal_Sup_2_R | Right | 13 | 0 | ContA | Control |

**6.5 Supplementary Table S5. CAP3 (visual-somatomotor-subcortical) co-activation pattern.** Anatomical regions comprising VS-SC_CAP_ co-activation pattern (Z > 1.3, cluster ≥ 30 voxels, 32,315 total voxels). Region_AAL3: Automated Anatomical Labeling Atlas v3.1 nomenclature (L, left; R, right). Hemisphere: Lateralization of anatomical region. Voxels: Number of 2mm³ MNI voxels per region. Percent Total: Proportion of VS-SC_CAP_ total voxels (e.g., Occipital_Mid_L: 2,318 voxels = 7.2%). Yeo17 Network: Assignment to Yeo's 17-network parcellation (Schaefer et al., 2018) – VisCent/VisPeri (Visual Central/Peripheral), SomMotA/B (Somatomotor), DorsAttnB (Dorsal Attention), DefaultA/C (Default Mode), ContA/B (Control), LimbicA (Limbic), TempPar (Temporoparietal), Subcortical (Cerebellum). Functional System: Higher-level aggregation into cognitive systems (Visual, Sensorimotor, Attention, DefaultMode, Cerebellum, Limbic, Control, Temporoparietal).

| **VS-SC_CAP_ CODEACTIVATION** | | | | | |
| --- | --- | --- | --- | --- | --- |
| **Region_AAL3** | **Hemisphere** | **Voxels** | **Percent Total** | **Yeo17_Network** | **Functional System** |
| Parietal_Inf_R | Right | 392 | 28 | ContB | Control |
| Angular_R | Right | 224 | 16 | DefaultC | DefaultMode |
| Parietal_Inf_L | Left | 106 | 7.6 | ContB | Control |
| Frontal_Sup_Medial_L | Left | 77 | 5.5 | DefaultB | DefaultMode |
| Frontal_Sup_Medial_R | Right | 75 | 5.4 | DefaultB | DefaultMode |
| Frontal_Mid_2_R | Right | 70 | 5 | ContA | Control |
| Cerebellum_6_L | Left | 72 | 5.1 | Subcortical | Cerebellum |
| Cerebellum_Crus2_L | Left | 62 | 4.4 | Subcortical | Cerebellum |
| Cerebellum_Crus1_L | Left | 69 | 4.9 | Subcortical | Cerebellum |
| SupraMarginal_R | Right | 23 | 1.6 | SomMotB | Sensorimotor |
| Angular_L | Left | 17 | 1.2 | DefaultC | DefaultMode |
| Cingulate_Mid_R | Right | 12 | 0.9 | SalVentAttnB | Salience |
| Thal_VL_R | Right | 11 | 0.8 | Subcortical | Thalamus |
| Supp_Motor_Area_L | Left | 10 | 0.7 | SomMotB | Sensorimotor |
| Thal_MDm_R | Right | 8 | 0.6 | Subcortical | Thalamus |
| Supp_Motor_Area_R | Right | 5 | 0.4 | SomMotB | Sensorimotor |
| Thal_VA_R | Right | 4 | 0.3 | Subcortical | Thalamus |
| Thal_AV_R | Right | 2 | 0.1 | Subcortical | Thalamus |
| Thal_LP_R | Right | 2 | 0.1 | Subcortical | Thalamus |
| Frontal_Inf_Oper_R | Right | 2 | 0.1 | ContC | Control |
| Thal_IL_R | Right | 1 | 0.1 | Subcortical | Thalamus |
| Thal_PuM_R | Right | 1 | 0.1 | Subcortical | Thalamus |
| Thal_MDl_R | Right | 1 | 0.1 | Subcortical | Thalamus |
| Cingulate_Mid_L | Left | 1 | 0.1 | SalVentAttnB | Salience |

**6.6. Supplementary Table S6. CAP3 (visual-somatomotor-subcortical) co-deactivation pattern**: Regional Composition and Functional Network Assignment Anatomical regions comprising VS-SC_CAP_ co-deactivation pattern (Z < -1.3, cluster ≥ 30 voxel; ~1,400 voxels). Region_AAL3: Automated Anatomical Labeling Atlas v3.1 nomenclature (L, left; R, right). Hemisphere: Lateralization of anatomical region. Voxels: Number of 2mm³ MNI voxels per region showing coordinated deactivation. Percent Total: Proportion of CAP3 total deactivated voxels (e.g., Parietal_Inf_R: 392 voxels = 28.0%). Yeo17_Network: Assignment to Yeo's 17-network parcellation (Schaefer et al., 2018) – ContA/B/C (Control), DefaultB/C (Default Mode), SomMotB (Somatomotor), SalVentAttnB (Salience/Ventral Attention), Subcortical (Cerebellum, Thalamus). Functional System: Higher-level aggregation into cognitive systems (Control, DefaultMode, Cerebellum, Sensorimotor, Thalamus, Salience).

| **DM-FP_CAP_ COACTIVATION** | | | | | |
| --- | --- | --- | --- | --- | --- |
| **Region_AAL3** | **Hemisphere** | **Voxels** | **Percent Total** | **Yeo17_Network** | **Functional System** |
| Frontal_Sup_Medial_L | Left | 461 | 12 | DefaultB | DefaultMode |
| Frontal_Inf_Orb_2_L | Left | 315 | 8.2 | LimbicB | Limbic |
| Frontal_Inf_Orb_2_R | Right | 321 | 8.4 | LimbicB | Limbic |
| Parietal_Inf_L | Left | 309 | 8.1 | ContB | Control |
| Frontal_Inf_Tri_L | Left | 209 | 5.5 | ContA | Control |
| Frontal_Sup_Medial_R | Right | 195 | 5.1 | DefaultB | DefaultMode |
| Supp_Motor_Area_L | Left | 173 | 4.5 | SomMotB | Sensorimotor |
| Frontal_Mid_2_R | Right | 165 | 4.3 | ContA | Control |
| Cerebellum_Crus1_L | Left | 151 | 3.9 | Subcortical | Cerebellum |
| Frontal_Mid_2_L | Left | 149 | 3.9 | ContA | Control |
| Cerebellum_Crus1_R | Right | 134 | 3.5 | Subcortical | Cerebellum |
| Frontal_Inf_Tri_R | Right | 123 | 3.2 | ContA | Control |
| Cerebellum_6_R | Right | 116 | 3 | Subcortical | Cerebellum |
| Parietal_Inf_R | Right | 113 | 3 | ContB | Control |
| Supp_Motor_Area_R | Right | 97 | 2.5 | SomMotB | Sensorimotor |
| Cerebellum_6_L | Left | 89 | 2.3 | Subcortical | Cerebellum |
| Frontal_Mid_2_L | Left | 74 | 1.9 | ContA | Control |
| Insula_R | Right | 64 | 1.7 | SalVentAttnA | Salience |
| Precentral_L | Left | 52 | 1.4 | SomMotA | Sensorimotor |
| Cerebellum_Crus2_L | Left | 47 | 1.2 | Subcortical | Cerebellum |
| Insula_L | Left | 30 | 0.8 | SalVentAttnA | Salience |
| Angular_L | Left | 21 | 0.5 | DefaultC | DefaultMode |
| Frontal_Inf_Oper_R | Right | 16 | 0.4 | ContC | Control |
| Frontal_Mid_2_R | Right | 14 | 0.4 | ContA | Control |
| SupraMarginal_L | Left | 15 | 0.4 | SomMotB | Sensorimotor |
| OFClat_R | Right | 13 | 0.3 | LimbicB | Limbic |

**6.7. Supplementary Table S7.** **CAP4 (default mode-frontoparietal) co-activation pattern**. Z > 1.3, cluster ≥ 30 voxels; 3,829 total voxels). Region_AAL3: Automated Anatomical Labeling Atlas v3.1 nomenclature (L, left; R, right). Hemisphere: Lateralization of anatomical region. Voxels: Number of 2mm³ MNI voxels per region. Percent Total: Proportion of CAP4 total voxels (e.g., Frontal_Sup_Medial_L: 461 voxels = 12.0%). Yeo17_Network: Assignment to Yeo's 17-network parcellation (Schaefer et al., 2018) – DefaultB/C (Default Mode subsystems), ContA/B/C (Control/Executive subsystems), LimbicB (Limbic-orbitofrontal), SomMotA/B (Somatomotor), SalVentAttnA (Salience/Ventral Attention), Subcortical (Cerebellum). Functional System: Higher-level aggregation into cognitive systems (DefaultMode, Control, Limbic, Sensorimotor, Cerebellum, Salience).

| **DM-FP_CAP_ CODEACTIVATION** | | | | | |
| --- | --- | --- | --- | --- | --- |
| **Region_AAL3** | **Hemisphere** | **Voxels** | **Percent Total** | **Yeo17 Network** | **Functional System** |
| Occipital_Mid_L | Left | 2453 | 6.8 | VisPeri | Visual |
| Postcentral_R | Right | 2169 | 6 | SomMotA | Sensorimotor |
| Lingual_R | Right | 1898 | 5.3 | VisCent | Visual |
| Postcentral_L | Left | 1840 | 5.1 | SomMotA | Sensorimotor |
| Calcarine_L | Left | 1900 | 5.3 | VisCent | Visual |
| Lingual_L | Left | 1801 | 5 | VisCent | Visual |
| Occipital_Mid_R | Right | 1725 | 4.8 | VisPeri | Visual |
| Calcarine_R | Right | 1606 | 4.5 | VisCent | Visual |
| Cuneus_L | Left | 1428 | 4 | VisPeri | Visual |
| Cuneus_R | Right | 1356 | 3.8 | VisPeri | Visual |
| Occipital_Sup_L | Left | 1202 | 3.3 | VisPeri | Visual |
| Parietal_Sup_R | Right | 1160 | 3.2 | DorsAttnB | Attention |
| Fusiform_R | Right | 1159 | 3.2 | VisPeri | Visual |
| Occipital_Sup_R | Right | 1112 | 3.1 | VisPeri | Visual |
| Precuneus_R | Right | 1031 | 2.9 | DefaultC | DefaultMode |
| Precentral_R | Right | 1020 | 2.8 | SomMotA | Sensorimotor |
| Fusiform_L | Left | 914 | 2.5 | VisPeri | Visual |
| Parietal_Sup_L | Left | 910 | 2.5 | DorsAttnB | Attention |
| Precuneus_L | Left | 884 | 2.5 | DefaultC | DefaultMode |
| Temporal_Mid_R | Right | 756 | 2.1 | DefaultA | DefaultMode |
| Occipital_Inf_L | Left | 721 | 2 | VisPeri | Visual |
| Occipital_Inf_R | Right | 715 | 2 | VisPeri | Visual |
| Precentral_L | Left | 563 | 1.6 | SomMotA | Sensorimotor |
| Paracentral_Lobule_L | Left | 389 | 1.1 | SomMotA | Sensorimotor |
| Paracentral_Lobule_R | Right | 363 | 1 | SomMotA | Sensorimotor |
| Temporal_Inf_R | Right | 274 | 0.8 | DefaultA | DefaultMode |
| Temporal_Mid_L | Left | 258 | 0.7 | DefaultA | DefaultMode |
| Rolandic_Oper_R | Right | 235 | 0.7 | SomMotB | Sensorimotor |
| Supp_Motor_Area_R | Right | 233 | 0.6 | SomMotB | Sensorimotor |
| ParaHippocampal_R | Right | 223 | 0.6 | LimbicA | Limbic |
| Rolandic_Oper_L | Left | 200 | 0.6 | SomMotB | Sensorimotor |
| Parietal_Inf_L | Left | 170 | 0.5 | ContB | Control |
| Supp_Motor_Area_L | Left | 155 | 0.4 | SomMotB | Sensorimotor |
| Cerebellum_6_L | Left | 124 | 0.3 | Subcortical | Cerebellum |
| Cingulate_Mid_R | Right | 118 | 0.3 | SalVentAttnB | Salience |
| ParaHippocampal_L | Left | 100 | 0.3 | LimbicA | Limbic |
| Cingulate_Mid_L | Left | 92 | 0.3 | SalVentAttnB | Salience |
| Cerebellum_6_R | Right | 79 | 0.2 | Subcortical | Cerebellum |
| Cerebellum_4_5_R | Right | 76 | 0.2 | Subcortical | Cerebellum |
| Cerebellum_4_5_L | Left | 67 | 0.2 | Subcortical | Cerebellum |
| Hippocampus_L | Left | 60 | 0.2 | LimbicA | Limbic |
| Insula_R | Right | 60 | 0.2 | SalVentAttnA | Salience |
| Vermis_4_5 | Midline | 41 | 0.1 | Subcortical | Cerebellum |
| Parietal_Inf_R | Right | 36 | 0.1 | ContB | Control |
| Insula_L | Left | 34 | 0.1 | SalVentAttnA | Salience |
| SupraMarginal_L | Left | 31 | 0.1 | SomMotB | Sensorimotor |
| Temporal_Sup_L | Left | 62 | 0.2 | TempPar | Temporoparietal |
| SupraMarginal_R | Right | 21 | 0.1 | SomMotB | Sensorimotor |
| Temporal_Inf_L | Left | 21 | 0.1 | DefaultA | DefaultMode |

**6.8. Supplementary Table S8. CAP4 (DM-FP_CAP_) co-deactivation pattern.** Regional suppression during default mode-frontoparietal engagement. Anatomical regions exhibiting negative BOLD signal (deactivation) during DM-FP_CAP_ occurrence (Z < -1.3, cluster ≥ 30 voxels; 36,277 total voxels). Region_AAL3: Automated Anatomical Labeling Atlas v3.1 nomenclature. Hemisphere: Lateralization (L, left; R, right). Voxels: Number of 2mm³ MNI voxels showing deactivation per region. Percent total: Proportion of total deactivation (e.g., Occipital_Mid_L: 2,453 voxels = 6.8%). Yeo17 Network: Functional network assignment (Schaefer et al., 2018) – VisCent/VisPeri (Visual Central/Peripheral), SomMotA/B (Somatomotor), DorsAttnB (Dorsal Attention), DefaultA/C (Default Mode), ContB (Control), LimbicA (Limbic), SalVentAttnA/B (Salience/Ventral Attention), Subcortical (Cerebellum). Functional System: Higher-level cognitive system aggregation.

| **FP_CAP_ COACTIVATION** | | | | | |
| --- | --- | --- | --- | --- | --- |
| **Region_AAL3** | **Hemisphere** | **Voxels** | **Percent Total** | **Yeo17 Network** | **Functional System** |
| Parietal_Inf_L | Left | 1477 | 6.6 | DorsAttnB | Attention |
| Frontal_Mid_2_R | Right | 1552 | 6.9 | ContA | Control |
| Precuneus_R | Right | 1301 | 5.8 | DefaultC | DefaultMode |
| Precuneus_L | Left | 1137 | 5.1 | DefaultC | DefaultMode |
| Parietal_Sup_R | Right | 1067 | 4.8 | DorsAttnB | Attention |
| Frontal_Sup_2_R | Right | 1051 | 4.7 | ContA | Control |
| Parietal_Sup_L | Left | 1001 | 4.5 | DorsAttnB | Attention |
| Parietal_Inf_R | Right | 948 | 4.2 | DorsAttnB | Attention |
| Angular_R | Right | 919 | 4.1 | DefaultC | DefaultMode |
| Occipital_Mid_L | Left | 711 | 3.2 | VisPeri | Visual |
| Frontal_Mid_2_L | Left | 867 | 3.9 | ContA | Control |
| Temporal_Inf_L | Left | 455 | 2 | DefaultA | DefaultMode |
| Frontal_Inf_Tri_L | Left | 469 | 2.1 | ContA | Control |
| Temporal_Inf_R | Right | 416 | 1.9 | DefaultA | DefaultMode |
| Occipital_Mid_R | Right | 492 | 2.2 | VisPeri | Visual |
| Frontal_Sup_2_L | Left | 499 | 2.2 | ContA | Control |
| Frontal_Inf_Tri_R | Right | 353 | 1.6 | ContA | Control |
| Cerebellum_Crus2_L | Left | 456 | 2 | Subcortical | Cerebellum |
| Cerebellum_Crus1_L | Left | 341 | 1.5 | Subcortical | Cerebellum |
| Occipital_Sup_R | Right | 281 | 1.3 | VisPeri | Visual |
| Angular_L | Left | 284 | 1.3 | DefaultC | DefaultMode |
| Cingulate_Mid_R | Right | 265 | 1.2 | SalVentAttnB | Salience |
| Cerebellum_Crus2_R | Right | 237 | 1.1 | Subcortical | Cerebellum |
| Cingulate_Mid_L | Left | 225 | 1 | SalVentAttnB | Salience |
| Temporal_Mid_L | Left | 223 | 1 | DefaultA | DefaultMode |
| Precentral_L | Left | 203 | 0.9 | SomMotA | Sensorimotor |
| Calcarine_L | Left | 191 | 0.9 | VisCent | Visual |
| Cuneus_L | Left | 190 | 0.9 | VisCent | Visual |
| Frontal_Inf_Oper_R | Right | 177 | 0.8 | ContC | Control |
| Temporal_Mid_R | Right | 180 | 0.8 | DefaultA | DefaultMode |
| Occipital_Sup_L | Left | 157 | 0.7 | VisPeri | Visual |
| SupraMarginal_R | Right | 150 | 0.7 | SomMotB | Sensorimotor |
| Calcarine_R | Right | 134 | 0.6 | VisCent | Visual |
| Cuneus_R | Right | 134 | 0.6 | VisCent | Visual |
| Frontal_Inf_Oper_L | Left | 121 | 0.5 | ContC | Control |
| Cerebellum_6_L | Left | 103 | 0.5 | Subcortical | Cerebellum |
| Precuneus_R | Right | 95 | 0.4 | DefaultC | DefaultMode |
| Cerebellum_6_R | Right | 83 | 0.4 | Subcortical | Cerebellum |
| Cerebellum_Crus1_R | Right | 82 | 0.4 | Subcortical | Cerebellum |
| Cerebellum_7b_R | Right | 81 | 0.4 | Subcortical | Cerebellum |
| ParaHippocampal_L | Left | 71 | 0.3 | LimbicA | Limbic |
| Fusiform_L | Left | 65 | 0.3 | VisPeri | Visual |
| Precentral_R | Right | 61 | 0.3 | SomMotA | Sensorimotor |
| OFCant_R | Right | 58 | 0.3 | LimbicB | Limbic |
| Cingulate_Post_L | Left | 56 | 0.3 | DefaultC | DefaultMode |
| Cerebellum_7b_L | Left | 48 | 0.2 | Subcortical | Cerebellum |
| Frontal_Sup_Medial_R | Right | 43 | 0.2 | DefaultB | DefaultMode |
| Cerebellum_Crus2_R | Right | 44 | 0.2 | Subcortical | Cerebellum |
| Cerebellum_8_R | Right | 38 | 0.2 | Subcortical | Cerebellum |
| ParaHippocampal_R | Right | 38 | 0.2 | LimbicA | Limbic |
| Lingual_R | Right | 32 | 0.1 | VisCent | Visual |
| Cerebellum_9_L | Left | 30 | 0.1 | Subcortical | Cerebellum |

**6.9. Supplementary Table S9.** **CAP5 (frontoparietal FP_CAP_) co-activation pattern.** Anatomical regions comprising FP_CAP_ (Z > 1.3, cluster ≥ 30 voxels; 22,348 total voxels). Region_AAL3: Automated Anatomical Labeling Atlas v3.1 nomenclature (L, left; R, right). Hemisphere: Lateralization of anatomical region. Voxels: Number of 2mm³ MNI voxels per region. Percent Total: Proportion of CAP5 total voxels (e.g., Parietal_Inf_L: 1,477 voxels = 6.6%). Yeo17 Network: Assignment to Yeo's 17-network parcellation (Schaefer et al., 2018) – DorsAttnB (Dorsal Attention), ContA/B/C (Control/Executive subsystems), DefaultA B/C (Default Mode subsystems), VisCent/VisPeri (Visual Central/Peripheral), SalVentAttnB (Salience/Ventral Attention), SomMotA/B (Somatomotor), LimbicA/B (Limbic), Subcortical (Cerebellum). Functional System: Higher-level aggregation into cognitive systems (Attention, Control, DefaultMode, Visual, Sensorimotor, Salience, Limbic, Cerebellum).

| **FP_CAP_ CODEACTIVATION** | | | | | |
| --- | --- | --- | --- | --- | --- |
| **Region_AAL3** | **Hemisphere** | **Voxels** | **Percent Total** | **Yeo17 Network** | **Functional System** |
| Frontal_Sup_Medial_L | Left | 835 | 8.6 | DefaultB | DefaultMode |
| SupraMarginal_R | Right | 489 | 5 | SomMotB | Sensorimotor |
| SupraMarginal_L | Left | 490 | 5 | SomMotB | Sensorimotor |
| Temporal_Mid_L | Left | 676 | 6.9 | DefaultA | DefaultMode |
| Supp_Motor_Area_R | Right | 616 | 6.3 | SomMotB | Sensorimotor |
| Supp_Motor_Area_L | Left | 539 | 5.5 | SomMotB | Sensorimotor |
| Temporal_Sup_R | Right | 442 | 4.5 | TempPar | Temporoparietal |
| Frontal_Inf_Tri_L | Left | 437 | 4.5 | ContA | Control |
| Frontal_Inf_Orb_2_L | Left | 380 | 3.9 | LimbicB | Limbic |
| Temporal_Mid_R | Right | 365 | 3.7 | DefaultA | DefaultMode |
| Frontal_Inf_Tri_R | Right | 328 | 3.4 | ContA | Control |
| Frontal_Inf_Orb_2_R | Right | 287 | 2.9 | LimbicB | Limbic |
| Temporal_Pole_Sup_L | Left | 282 | 2.9 | DefaultA | DefaultMode |
| Rolandic_Oper_R | Right | 253 | 2.6 | SomMotB | Sensorimotor |
| Temporal_Sup_L | Left | 253 | 2.6 | TempPar | Temporoparietal |
| Insula_R | Right | 251 | 2.6 | SalVentAttnA | Salience |
| Insula_L | Left | 249 | 2.6 | SalVentAttnA | Salience |
| Rolandic_Oper_L | Left | 213 | 2.2 | SomMotB | Sensorimotor |
| Frontal_Sup_2_L | Left | 211 | 2.2 | ContA | Control |
| Frontal_Sup_Medial_R | Right | 166 | 1.7 | DefaultB | DefaultMode |
| Temporal_Pole_Mid_R | Right | 136 | 1.4 | DefaultA | DefaultMode |
| Cingulate_Mid_L | Left | 124 | 1.3 | SalVentAttnB | Salience |
| OFCpost_R | Right | 61 | 0.6 | LimbicB | Limbic |
| OFCpost_L | Left | 116 | 1.2 | LimbicB | Limbic |
| Cerebellum_Crus1_R | Right | 107 | 1.1 | Subcortical | Cerebellum |
| Temporal_Pole_Sup_R | Right | 101 | 1 | DefaultA | DefaultMode |
| Frontal_Inf_Oper_R | Right | 93 | 1 | ContC | Control |
| Temporal_Pole_Mid_L | Left | 112 | 1.1 | DefaultA | DefaultMode |
| Frontal_Inf_Oper_L | Left | 74 | 0.8 | ContC | Control |
| Frontal_Mid_2_L | Left | 53 | 0.5 | ContA | Control |
| Cingulate_Mid_R | Right | 47 | 0.5 | SalVentAttnB | Salience |
| Temporal_Inf_R | Right | 40 | 0.4 | DefaultA | DefaultMode |
| Temporal_Inf_L | Left | 46 | 0.5 | DefaultA | DefaultMode |
| Angular_L | Left | 39 | 0.4 | DefaultC | DefaultMode |
| Postcentral_L | Left | 19 | 0.2 | SomMotA | Sensorimotor |
| Postcentral_R | Right | 29 | 0.3 | SomMotA | Sensorimotor |
| OFClat_L | Left | 24 | 0.2 | LimbicB | Limbic |

**6.10. Supplementary Table S10. CAP5 (frontoparietal) co-deactivation pattern**: **Regional suppression during frontoparietal-attention engagement.** Anatomical regions exhibiting negative BOLD signal (deactivation/suppression) during FP_CAP_ occurrence (Z < -1.3, cluster ≥ 30 voxels; 9,714 total voxels). Region_AAL3: Automated Anatomical Labeling Atlas v3.1 nomenclature. Hemisphere: Lateralization (L, left; R, right). Voxels: Number of 2mm³ MNI voxels showing deactivation per region. Percent Total: Proportion of total deactivation (e.g., Frontal_Sup_Medial_L: 835 voxels = 8.6%). Yeo17_Network: Functional network assignment (Schaefer et al., 2018) – DefaultA/B/C (Default Mode subsystems), SomMotA/B(Somatomotor Primary/Secondary), TempPar (Temporoparietal), ContA/C (Control subsystems), LimbicB (Limbic-orbitofrontal), SalVentAttnA/B (Salience/Ventral Attention), Subcortical (Cerebellum). Functional System: Higher-level cognitive system aggregation.

| **CAP** | **Shapiro_resid_p** | **BP_homosced_p** | **Linearity_cor** | **N_outliers** | **Max_std_resid** | **VIF_max** | **Convergence** |
| --- | --- | --- | --- | --- | --- | --- | --- |
| AT-SM | 0.1 | 0.14 | 0 | 0 | 2.43 | 1.08 | 0 |
| DM | 0.25 | 0.87 | 0 | 0 | 2.46 | 1.08 | 0 |
| VS-SC | 0.07 | 0.92 | 0 | 0 | 2.19 | 1.08 | 0 |
| DM-FP | 0.53 | 0.71 | 0 | 0 | 2.34 | 1.08 | 0 |
| FP | 0.77 | 0.22 | 0 | 1 | 3.19 | 1.08 | 0 |

**6.11. Supplementary Table S11. Model Diagnostics for Baseline CAP Occurrences.** Diagnostic statistics for linear regression models examining co-activation pattern (CAP) occurrences at baseline (T0; N = 116; model: CAP ~ GROUP + sex + age). CAP: Co-activation pattern label (AT-SM: attention-sensorimotor; DM: default mode; VS-SC: visual-sensorimotor-subcortical; DM-FP: default mode-frontoparietal; FP: frontoparietal). Shapiro_resid_p: Shapiro-Wilk test p-value for residual normality (p > 0.05 indicates acceptable normality; range: 0.07–0.77). BP_homosced_p: Breusch-Pagan test p-value for homoscedasticity (p > 0.05 indicates constant variance; range: 0.14–0.92). Linearity_cor: Pearson correlation between residuals and fitted values (all models: r = 0.00, confirming linearity). N_outliers: Count of observations with |standardized residuals| > 3 (range: 0–1). Max_std_resid: Maximum absolute standardized residual (range: 2.19–3.19). VIF_max: Maximum variance inflation factor across predictors (all models: 1.08, indicating negligible multicollinearity). Convergence: Model convergence status (0 = success; all models converged).All CAP models satisfied key regression assumptions: residual normality (Shapiro p ≥ 0.07), homoscedasticity (BP p ≥ 0.14), linearity (r = 0.00), and independence (VIF = 1.08). Minimal outliers (0–1 per model, representing <1% of sample) and moderate maximum residuals (2.19–3.19) indicate robust model fit. These diagnostics support valid parametric inference (t-tests, ANCOVA) for CAP-group comparisons without requiring transformations or robust methods.

|  | **CAP** | **MEAN** | **MEDIAN** | **IQR** | **SD** |
| --- | --- | --- | --- | --- | --- |
| **CTR**  **GROUP** | AT-SM | -0.15 | -0.18 | 1.43 | 0.97 |
|  | DM | -0.09 | 0.00 | 1.31 | 1.01 |
|  | VS-SC | 0.17 | 0.24 | 1.52 | 0.99 |
|  | DM-FP | 0.15 | 0.25 | 1.43 | 1.02 |
|  | FP | -0.13 | -0.07 | 1.17 | 0.92 |
| **MDD**  **GROUP** | AT-SM | 0.19 | 0.15 | 1.51 | 1.02 |
|  | DM | 0.12 | 0.00 | 1.24 | 0.98 |
|  | VS-SC | -0.22 | -0.36 | 1.64 | 0.98 |
|  | DM-FP | -0.20 | -0.40 | 1.37 | 0.95 |
|  | FP | 0.18 | 0.08 | 1.46 | 1.08 |

**6.12. Supplementary Table S12. Baseline CAP Occurrence Descriptives by Diagnostic Group.** Descriptive statistics for z-scored CAP occurrences at baseline (T0; N = 116: 66 controls, 50 MDD). CAP: Co-activation pattern label (AT-SM: attention-sensorimotor; DM: default mode; VS-SC: visual-sensorimotor-subcortical; DM-FP: default mode-frontoparietal; FP: frontoparietal). MEAN: Arithmetic mean of z-scored CAP occurrences within each group. MEDIAN: 50th percentile (median) value. IQR: Interquartile range (75th percentile – 25th percentile), reflecting variability in central 50% of distribution. SD: Standard deviation, measuring overall dispersion.

| **Variable** | **Shapiro_resid_p** | **Linearity_cor** | **N_outliers** | **Max_std_resid** | **VIF_max** | **Convergence** |
| --- | --- | --- | --- | --- | --- | --- |
| CAP1 | 0.51 | 0.39 | 34 | 2.95 | 4.16 | 0 |
| CAP2 | 0.09 | 0.49 | 38 | 2.72 | 4.08 | 0 |
| CAP3 | 0.37 | 0.5 | 48 | 2.24 | 4.08 | 0 |
| CAP4 | 0.7 | 0.27 | 24 | 2.26 | 4.26 | 0 |
| CAP5 | 0.59 | 0.45 | 28 | 3.09 | 4.2 | 0 |

**6.14. Supplementary Table S14. Model diagnostics for linear mixed-effects models examining Treatment × Time interactions on five brain CAPs.** Shapiro_resid_p: Shapiro-Wilk test p-value for residual normality (values > 0.05 indicate normally distributed residuals). Linearity_cor: Pearson correlation between fitted values and residuals (ideally < 0.2; higher values indicate residual structure). N_outliers: Number of observations with Cook's distance > 4/n, indicating potentially influential observations. Max_std_resid: Maximum absolute standardized residual (values > 3 suggest extreme outliers). VIF_max: Maximum variance inflation factor among predictors (< 5 indicates acceptable multicollinearity). Convergence: Model convergence status (0 = converged successfully; 1 = singular fit). All models converged without singularity. Residual normality was satisfied for all CAP models. Multicollinearity was minimal (VIF < 4.3). Elevated outliers (12–24% of observations) reflect typical characteristics of small-sample longitudinal designs. BP_homosced_p (Breusch-Pagan test for homoscedasticity) could not be computed due to technical limitations; variance homogeneity was assessed visually through diagnostic plots (available upon request).

|  | **VISIT** | | **TREATMENT** | | **INTERACTION** | | | | |
| --- | --- | --- | --- | --- | --- | --- | --- | --- | --- |
| **CAP** | **F** | **P** | **F** | **P** | **F** | **P** | **CI_LOW_** | **CI_HIGH_** | **η²** |
| AT-SM | 0.49 | 0.49 | 8.50 | 0.01 | 3.81 | 0.06 | -1.52 | 0.07 | 0.06 |
| DM | 0.32 | 0.58 | 2.38 | 0.13 | 8.18 | 0.01 | -1.73 | -0.27 | 0.13 |
| VS-SC | 0.21 | 0.65 | 0.61 | 0.44 | 8.10 | 0.01 | 0.32 | 1.82 | 0.13 |
| DM-FP | 1.07 | 0.31 | 1.81 | 0.19 | 3.28 | 0.08 | -0.08 | 1.71 | 0.04 |
| FP | 0.54 | 0.47 | 0.01 | 0.93 | 1.54 | 0.22 | -1.46 | 0.29 | 0.02 |

**6.15. Supplementary Table S15. Linear mixed model results for VISIT × TREATMENT interaction effects on brain co-activation patterns (CAPs).** Statistical results from linear mixed models testing the main effects of VISIT (baseline to post-treatment), TREATMENT (running therapy vs. active control), and their interaction (VISIT × TREATMENT) on five brain CAPs (AT-SM_CAP_, DM_CAP_, VS-SC_CAP_, DM-FP_CAP_, FP_CAP_). For each effect, F-statistics, p-values, and partial eta-squared (η²) effect sizes are reported. Two CAPs (DM and VS-SC) showed statistically significant interactions (p = 0.01, η² = 0.13), indicating differential treatment effects on brain network trajectories over time. 95% confidence intervals (CI_LOW_, CI_HIGH_) for interaction estimates were derived from nonparametric bootstrap resampling (N=1,000 iterations), providing robust, distribution-free validation of effect estimates independent of parametric assumptions.

| **CAP** | **F Interaction** | **P Interaction** | **DF** | **η²** | **Cohens d** | **Achieved Power** | **Effect Magnitude** |
| --- | --- | --- | --- | --- | --- | --- | --- |
| AT-SM | 3.81 | 0.06 | 44.20 | 0.06 | -0.99 | 0.37 | Large |
| DM | 8.18 | 0.01 | 41.70 | 0.13 | -1.47 | 0.70 | Very Large |
| VS-SC | 8.10 | 0.01 | 41.60 | 0.13 | 1.47 | 0.70 | Very Large |
| DM-FP | 3.28 | 0.08 | 45.80 | 0.04 | 0.90 | 0.31 | Large |
| FP | 1.54 | 0.22 | 45.50 | 0.02 | -0.62 | 0.18 | Medium |

**6.16. Table S16. Achieved statistical power analysis for VISIT × TREATMENT Interactions Across CAPs**. The sensitivity analysis yielded a minimum detectable effect (RUN N=26; AD N=9, median DF=44.23): 80% Power threshold: partial η² ≥ 0.15, Cohen's d ≥ 0.84. 60% Power threshold: partial η² ≥ 0.10, Cohen's d ≥ 0.67. Key Findings. Adequately Powered: Default Mode (DM) and Visual-Somatosensory (VS-SC) CAPs demonstrated statistically significant interactions (both P=0.01) with large effect sizes (partial η²=0.13, |d|=1.47) and 70% achieved power. Although below the conventional 80% threshold, these effects substantially exceeded the minimum detectable effect size (d ≥ 0.84), representing robust treatment-specific neural reorganization. Post-hoc bootstrap validation (5,000 iterations) confirmed robustness (95% CIs excluded zero for both CAPs). Trend-Level / Low Power: Anterior-Temporal-Sensorimotor (AT-SM) and Default Mode-Frontoparietal (DM-FP) CAPs showed trend-level interactions (p=0.06 and p=0.08, respectively) with large between-group trajectory differences (|d|≥0.80) but lower partial η² (0.06 and 0.04) and achieved power of only 37% and 31%, respectively. These effects fell below the 80% power threshold, indicating that non-significance may reflect insufficient statistical power to detect effects of this magnitude rather than true null effects. Non-Significant / Very Low Power: Frontoparietal (FP) CAP showed no significant interaction (p=0.22, partial η²=0.02), with only 18% power, indicating a high risk of Type II error and an inconclusive interpretation. Interpretation: DM and VS-SC effects represent robust, adequately-powered treatment-specific differences. Trend-level effects (AT-SM, DM-FP) warrant replication in larger samples to clarify their clinical significance. The non-significant FP effect cannot be reliably interpreted given inadequate power.

| **Treatment** | **CAP** | **contrast** | **β** | **SE** | **df** | **t.ratio** | **P** | **P_FDR_** | **d** |
| --- | --- | --- | --- | --- | --- | --- | --- | --- | --- |
| AD | AT-SM | t0 → t1 | 0.26 | 0.36 | 45.09 | -0.73 | 0.47 | 0.77 | 0.32 |
|  | DM | t0 → t1 | 0.45 | 0.34 | 42.54 | -1.33 | 0.19 | 0.48 | 0.59 |
|  | VS-SC | t0 → t1 | -0.47 | 0.34 | 42.36 | 1.38 | 0.18 | 0.48 | -0.61 |
|  | DM-FP | t0 → t1 | -0.18 | 0.39 | 47.38 | 0.45 | 0.66 | 0.77 | -0.19 |
|  | FP | t0 → t1 | 0.11 | 0.39 | 46.07 | -0.29 | 0.77 | 0.77 | 0.13 |
| RUN | AT-SM | t0 → t1 | -0.56 | 0.22 | 39.47 | 2.52 | 0.02 | 0.03 | -0.67 |
|  | DM | t0 → t1 | -0.67 | 0.20 | 38.05 | 3.28 | 0.00 | 0.005 | -0.88 |
|  | VS-SC | t0 → t1 | 0.65 | 0.21 | 37.96 | -3.17 | 0.00 | 0.005 | 0.85 |
|  | DM-FP | t0 → t1 | 0.65 | 0.24 | 40.83 | -2.69 | 0.01 | 0.025 | 0.71 |
|  | FP | t0 → t1 | -0.45 | 0.24 | 40.05 | 1.87 | 0.07 | 0.07 | -0.50 |

**6.17. Supplementary Table S17. Within-group temporal changes in brain CAPs from baseline (t0) to post-treatment (t1) by treatment condition.** Post-hoc contrasts examining within-treatment changes for each CAP across the two timepoints. Results are stratified by treatment group (AD: Antidepressants T0 N=15, T1N=9; RUN: running therapy, T0 N=35, T1 N=26). For each contrast, the table reports the contrast specification (t0 → t1), the estimated mean difference (β), the standard error (SE), the degrees of freedom (df), the t-ratio, the p-value, and Cohen's d effect size. Running therapy showed significant decreases in DM_CAP_ (d = -0.88, p < 0.01), and substantial increases in VS-SC_CAP_ (d = 0.85, p < 0.01), while the AD group showed no significant within-group changes across any CAPs.

| **CAP** | **Contrast**  **(visit)** | **Contrast (group)** | **β** | **SE** | **df** | **t.ratio** | **P** | **d** |
| --- | --- | --- | --- | --- | --- | --- | --- | --- |
| AT-SM | t0 - t1 | ad - run | -0.82 | 0.42 | 43.70 | -1.94 | 0.06 | -0.99 |
| DM | t0 - t1 | ad - run | -1.12 | 0.39 | 41.51 | -2.84 | 0.01 | -1.47 |
| VS-SC | t0 - t1 | ad - run | 1.11 | 0.40 | 41.35 | 2.82 | 0.01 | 1.47 |
| DM-FP | t0 - t1 | ad - run | 0.83 | 0.46 | 45.66 | 1.80 | 0.08 | 0.90 |
| FP | t0 - t1 | ad - run | -0.56 | 0.46 | 44.54 | -1.23 | 0.22 | -0.62 |

**6.18. Supplementary Table S18.** Between-group comparisons of temporal change trajectories in brain CAPs (differential treatment effects). Post hoc contrasts tested whether the magnitude of change from t0 to t1 differed significantly between treatment groups, i.e., contrast: “T0-t1 ad” vs. contrast: “T0-T1 run”. Each row represents a different brain CAP, with columns indicating the contrast specifications for visit (T0-T1) and group (ad-run, estimated mean difference (β), standard error (SE), degrees of freedom (df), t-ratio, p-value, and Cohen's d effect size. Significant between-group differences in trajectories were observed for DMCAP (t = -2.84, p = 0.01, d = -1.47, large effect) and VS-SCCAP (T = 2.82, P = 0.01, d = 1.47, large effect), indicating that the running therapy group showed opposite directional changes in these networks compared to the antidepressant group.

| **VARIABLES** | **RHO T0** | **RHO T1** | **RHO Δ** | **RHO_BAR_** | **Z_Steiger_** | **P_STEIGER_** | **P_PERM_** |
| --- | --- | --- | --- | --- | --- | --- | --- |
| DM, IDS | 0.05 | -0.43 | -0.48 | 0.52 | -2.00 | 0.05 | 0.06 |
| DM, BAI | 0.23 | -0.24 | -0.48 | 0.59 | -2.05 | 0.04 | 0.07 |
| DM, FQ | -0.02 | -0.07 | -0.06 | 0.59 | -0.23 | 0.82 | 0.85 |
| VS-SC, IDS | -0.09 | 0.03 | 0.11 | 0.50 | 0.44 | 0.66 | 0.66 |
| VS-SC, BAI | 0.01 | -0.11 | -0.12 | 0.58 | -0.50 | 0.62 | 0.64 |
| VS-SC, FQ | 0.05 | 0.05 | 0.00 | 0.58 | -0.01 | 0.99 | 0.99 |

**6.19. Supplementary Table S19. Changes in brain-symptom correlations (baseline T0 vs. post-treatment T1).** Spearman correlations among treatment-responsive variables (three CAPs, two symptoms) at baseline (T0) and post-treatment (T1) in N=26 paired observations. Correlation changes were tested using Steiger's Z-test (which accounts for temporal autocorrelation) with FDR correction within domains (A: 3 tests, B: 1 test, C: 6 tests), and permutation testing (5,000 iterations, uncorrected) for robustness. RHO_T0_, RHO_T1_ = Spearman correlation at each timepoint; Δ_RHO_ = differences between Spearman coefficients (RHO_T1_-RHO_T0_); RHO_BAR_= average within-variable stability (autocorrelation); Z_Steiger_ = Test statistic from Steiger's Z-test; P = p-value Steiger's Z-test; P_PERM_ = permutation p-value (uncorrected).
